# Refining Cardiovascular-Kidney-Metabolic Risk Assessment Across the Heart Failure Spectrum

**DOI:** 10.64898/2026.11.18.26368376

**Authors:** Su A Noh, Nan Young Bae, Jaehyun Lim, Soongu Kwak, Minjung Bak, Jiesuck Park, Hong-Mi Choi, Jun-Bean Park, Yeonyee E. Yoon, Seung-Pyo Lee, Jae-Hyeong Park, Yong-Jin Kim, Goo-Yeong Cho, Hyung-Kwan Kim, In-Chang Hwang

## Abstract

**Background:** The cardiovascular-kidney-metabolic (CKM) framework emphasizes risk assessment and prevention of heart failure (HF). However, the prognostic implications and optimal assessment of CKM burden in established HF remain uncertain.

**Objectives:** To evaluate the prognostic value of binary and HF-adapted graded CKM burden scores across HF phenotypes.

**Methods:** We evaluated 2 independent HF cohorts comprising patients with HF with reduced ejection fraction (HFrEF) initiating angiotensin receptor–neprilysin inhibitor therapy (HFrEF– ARNI; n=1,450) and patients hospitalized for acute HF (AHF; n = 2,985). AHF patients were stratified by left ventricular ejection fraction (LVEF) into <50% and ≥50% (HFrEF and HF with preserved ejection fraction [HFpEF] groups, respectively). The binary score counted 6 CKM components; the graded score incorporated chronic kidney disease severity and reverse-scored body mass index categories. Associations with the composite outcome of all-cause death or HF hospitalization were assessed using Cox regression. Prognostic discrimination and interactions with HF phenotype were also evaluated.

**Results:** Median follow-up was 31.5 months in HFrEF–ARNI and 51.1 months in AHF. Both scores were independently associated with the composite outcome in both cohorts. In HFrEF– ARNI cohort, adjusted HRs per 1-point increase were 1.10 (95% CI 1.02–1.18) for the binary score and 1.11 (95% CI 1.04–1.19) for the graded score. In AHF, both scores were associated with outcomes in HFrEF, whereas only the graded score was significantly associated with the outcome in HFpEF (binary: HR 0.96, 95% CI 0.90–1.02; graded: HR 1.09, 95% CI 1.04–1.15). Significant interactions with HF phenotype were observed for both scores. The graded score showed better discrimination than the binary score in both cohorts (C-index 0.632 vs 0.583 in HFrEF–ARNI; 0.609 vs 0.541 in AHF), particularly in HFpEF (C-index 0.576 vs 0.491). In HFpEF, only the graded score provided significant incremental prognostic information beyond clinical and echocardiographic variables (Δχ² = 11.3; P <0.001).

**Conclusions:** The HF-adapted graded CKM burden score demonstrated more consistent prognostic associations and better risk discrimination than the binary score, particularly in HFpEF. Incorporating disease severity and HF-relevant prognostic characteristics may refine CKM burden assessment and risk stratification in established HF.

**Condensed Abstract:** Cardiovascular-kidney-metabolic (CKM) burden is relevant to both heart failure (HF) development and prognosis in established HF. Previous studies used binary component counts, potentially overlooking differences in severity and prognostic implications. In two HF cohorts encompassing the full HF spectrum, we compared binary and HF-adapted graded scores incorporating chronic kidney disease severity and reverse-scored body mass index. The graded score showed more consistent associations with all-cause death or HF hospitalization and better discrimination, particularly in HF with preserved ejection fraction. Incorporating disease severity and HF-relevant prognostic characteristics may refine CKM burden assessment and risk stratification in established HF.

**Graphical Abstract:** 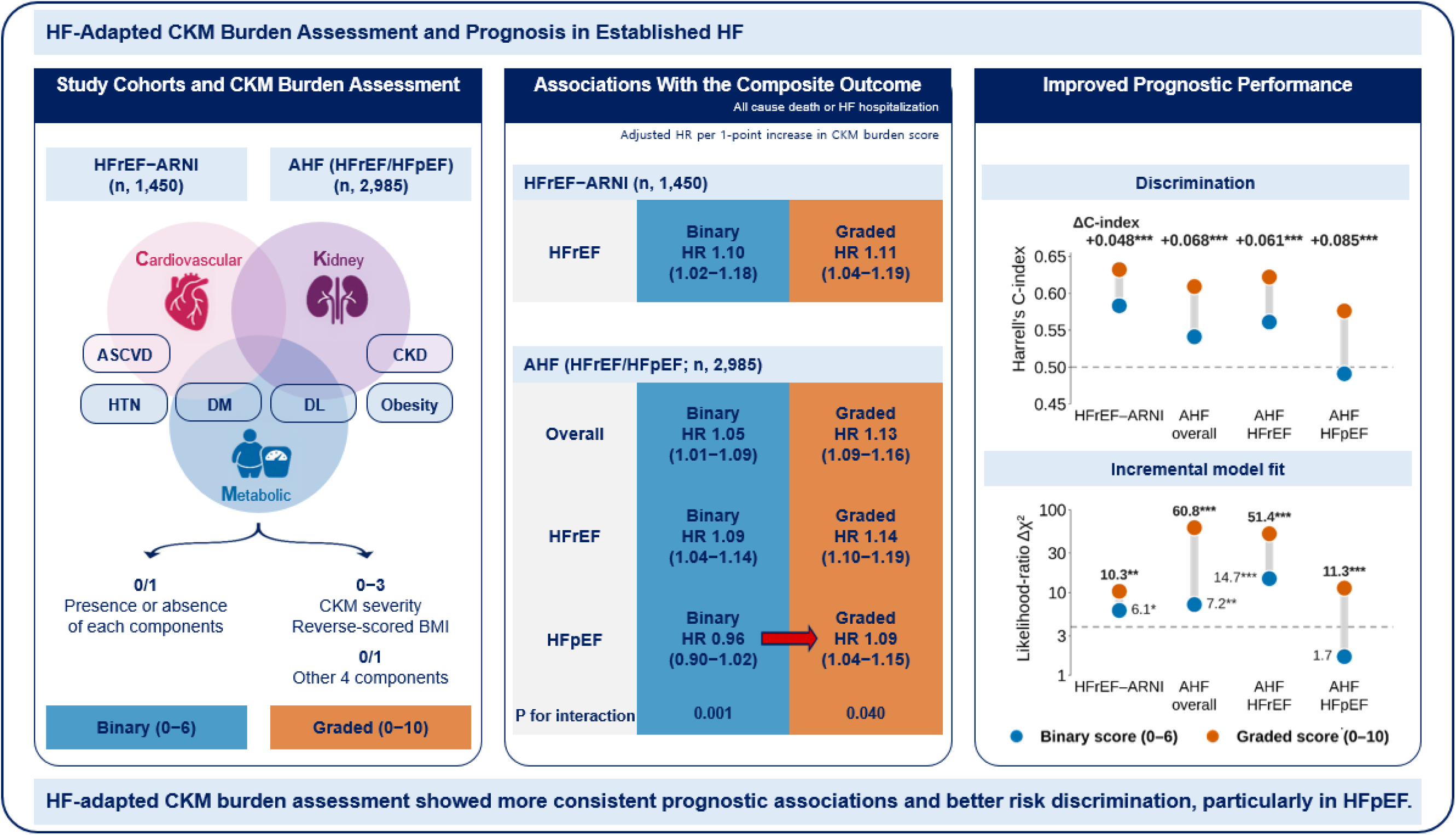

Binary and graded CKM burden scores were evaluated in the HFrEF–ARNI and AHF cohorts. The binary score captured the presence of six CKM components, whereas the graded score additionally incorporated reverse-scored BMI and CKD severity. The graded score demonstrated more consistent associations with the composite outcome of all-cause death or HF hospitalization, particularly in HFpEF. It also provided better prognostic performance across cohorts and HF phenotypes. Adjusted hazard ratios were estimated using multivariable Cox regression. Prognostic performance was assessed using Harrell’s C-index and likelihood-ratio χ²statistics. * *P* <0.05; ** *P* <0.01; *** *P* <0.001.

## Introduction

The cardiovascular-kidney-metabolic (CKM) syndrome framework has broadened cardiovascular risk assessment to explicitly encompass heart failure (HF) alongside atherosclerotic cardiovascular disease.^1^ Recognizing the interconnected contributions of metabolic abnormalities, kidney dysfunction, and cardiovascular risk factors, current guidance emphasizes integrated CKM assessment and management primarily for the prevention of cardiovascular disease, including HF. However, CKM components remain prevalent after HF develops and may continue to influence its clinical course.^2–7^ Although established HF with metabolic risk factors or chronic kidney disease (CKD) is classified as stage 4 CKM syndrome, this classification does not distinguish differences in CKM burden among patients with HF.^1, 3–7^ Assessing this burden may therefore help refine prognosis in established HF.

Although individual CKM components have prognostic value in HF, their cumulative burden may provide a more integrated assessment of conditions that influence clinical outcomes. Previous studies have examined CKM burden in patients with HF, commonly using binary definitions for each component.^3, 8^ However, counting CKM conditions does not account for differences in disease severity or their potentially distinct prognostic implications in established HF. An assessment incorporating these characteristics may therefore better characterize CKM-related risk. Whether this approach improves prognostic assessment compared with a binary component count, and whether its value differs across HF phenotypes, remains uncertain.

We therefore evaluated the prognostic implications of binary and HF-adapted graded CKM burden scores, which incorporated chronic kidney disease (CKD) severity and reverse-scored body mass index (BMI) categories, in two independent cohorts encompassing the full HF spectrum. We compared their associations with clinical outcomes and their prognostic discrimination, both overall and according to HF phenotype. We further examined associations of individual CKM components with clinical outcomes, including potential interactions with HF phenotype.

## Method

### Study Population

Clinical data were retrospectively obtained from two multicenter registries: STrain for Risk Assessment and Therapeutic Strategies in patients with Heart Failure treated with Angiotensin Receptor-Neprilysin Inhibitor (STRATS-HF-ARNI), which enrolled 2,757 consecutive patients with HF with reduced ejection fraction (HFrEF) initiating angiotensin receptor-neprilysin inhibitor therapy between 2017 and 2022,^9–12^ and STrain for Risk Assessment and Therapeutic Strategies in patients with Acute Heart Failure (STRATS-AHF), which enrolled 4,312 consecutive patients hospitalized for AHF between 2009 and 2016.^13–16^ Details of the registries have been published previously.^9–16^

For the present analysis, patients with complete data for all six CKM burden score components, left ventricular ejection fraction (LVEF), and the prespecified model covariates (age, sex, left ventricular global longitudinal strain [LVGLS], left atrial reservoir strain [LASr], and natriuretic peptide levels) were included. The final study population comprised 1,450 patients from STRATS-HF-ARNI (HFrEF-ARNI cohort) and 2,985 patients from STRATS-AHF (AHF cohort). In the AHF cohort, patients were further stratified by LVEF into two HF phenotypes: HFrEF (LVEF <50%) and HF with preserved ejection fraction (HFpEF; LVEF ≥50%) for subgroup analyses.^17^ The study protocol was approved by the Institutional Review Boards at each center, and the requirement for informed consent was waived due to the retrospective study design.

### CKM burden scoring

A binary CKM burden score (0–6) was constructed using six CKM components available in both cohorts: hypertension, diabetes mellitus, atherosclerotic cardiovascular disease, dyslipidemia, obesity, and CKD, with 1 point assigned to the presence of each component. An HF-adapted graded CKM burden score (0–10) was developed based on component-level analyses in both cohorts. Specifically, CKD severity and BMI were each assigned 0–3 points according to estimated glomerular filtration rate (eGFR) and BMI categories, respectively, with higher scores reflecting more severe kidney dysfunction and lower BMI. This approach was based on the observed positive association between CKD severity and adverse outcomes and the inverse association between BMI and adverse outcomes (**Supplemental Figure 1**). The remaining four components retained binary scoring.

Both scores were analyzed continuously (per 1-point increment) and categorically (binary: 0–1, 2–3, and ≥4; graded: 0–2, 3–5, and ≥6). Category boundaries were selected based on observed risk gradients and category sizes, with identical cutoffs applied in both cohorts. Detailed scoring criteria and category selection methods are provided in **Supplemental Method 1**.

### Outcomes

The primary outcome was a composite of all-cause mortality and HF hospitalization. Secondary outcomes were the individual components of the composite outcome. Outcomes were ascertained through medical records and linkage with national mortality data.^9–13^ Follow-up was calculated from the index date, defined as the initiation of angiotensin receptor–neprilysin inhibitor therapy in the HFrEF-ARNI cohort and the index hospitalization in the AHF cohort.

### Statistical Analysis

Continuous variables were summarized as mean ± standard deviation or median (interquartile range), as appropriate, and categorical variables as counts and percentages. Comparisons between cohorts and between HF phenotypes within the AHF cohort were performed using Welch’s *t* test, the Mann–Whitney U test, or the χ² test as appropriate. Standardized mean differences were calculated to quantify between-group differences.

Time-to-event associations were assessed using Cox proportional hazards models adjusted for age, sex, LVGLS, LASr, and natriuretic peptide decile, separately for each cohort and each HF phenotype within the AHF cohort. LVGLS was expressed as the absolute value for ease in interpretation. Natriuretic peptide deciles were defined using previously published cutoffs for B-type natriuretic peptide and N-terminal pro-B-type natriuretic peptide (**Supplemental Table 1**).^13^ Unadjusted absolute risks were estimated using the Kaplan–Meier method, and incidence rates were expressed per 100 person-years. Competing-risk analyses performed for HF hospitalization. Discrimination was assessed using Harrell’s C-index for each score alone, and incremental prognostic value was evaluated using likelihood-ratio tests.

Individual CKM components were evaluated in separate adjusted Cox models, with differences between the HFrEF and HFpEF subgroups within the AHF cohort assessed using likelihood-ratio tests for component-by-phenotype interactions. Exploratory conditional associations among CKM components, echocardiographic measures, and clinical variables were examined using mixed graphical models with L1 regularization.

Analyses were based on complete cases, with outcome-specific inclusion determined by available follow-up data. All tests were two-sided, and P <0.05 indicated statistical significance. Analyses were performed using Python version 3.12 with the *lifelines*, *statsmodels*, *scikit-learn*, and *scipy* packages. Additional details of the prognostic and network analyses are provided in the **Supplemental Method 2**.

## Results

### Baseline Characteristics

Compared with the HFrEF-ARNI cohort, patients in the AHF cohort were older, more often female, and had higher natriuretic peptide deciles (**Table 1**). Hypertension, diabetes, dyslipidemia, and CKD were more prevalent in the AHF cohort, the prevalence of atherosclerotic cardiovascular disease was comparable and obesity was less prevalent. Patients in the HFrEF-ARNI cohort had lower LVEF and LVGLS, whereas LASr did not differ significantly between cohorts. Baseline characteristics by HF phenotype within the AHF cohort are summarized in **Supplemental Table 2**.

**Table 1.**
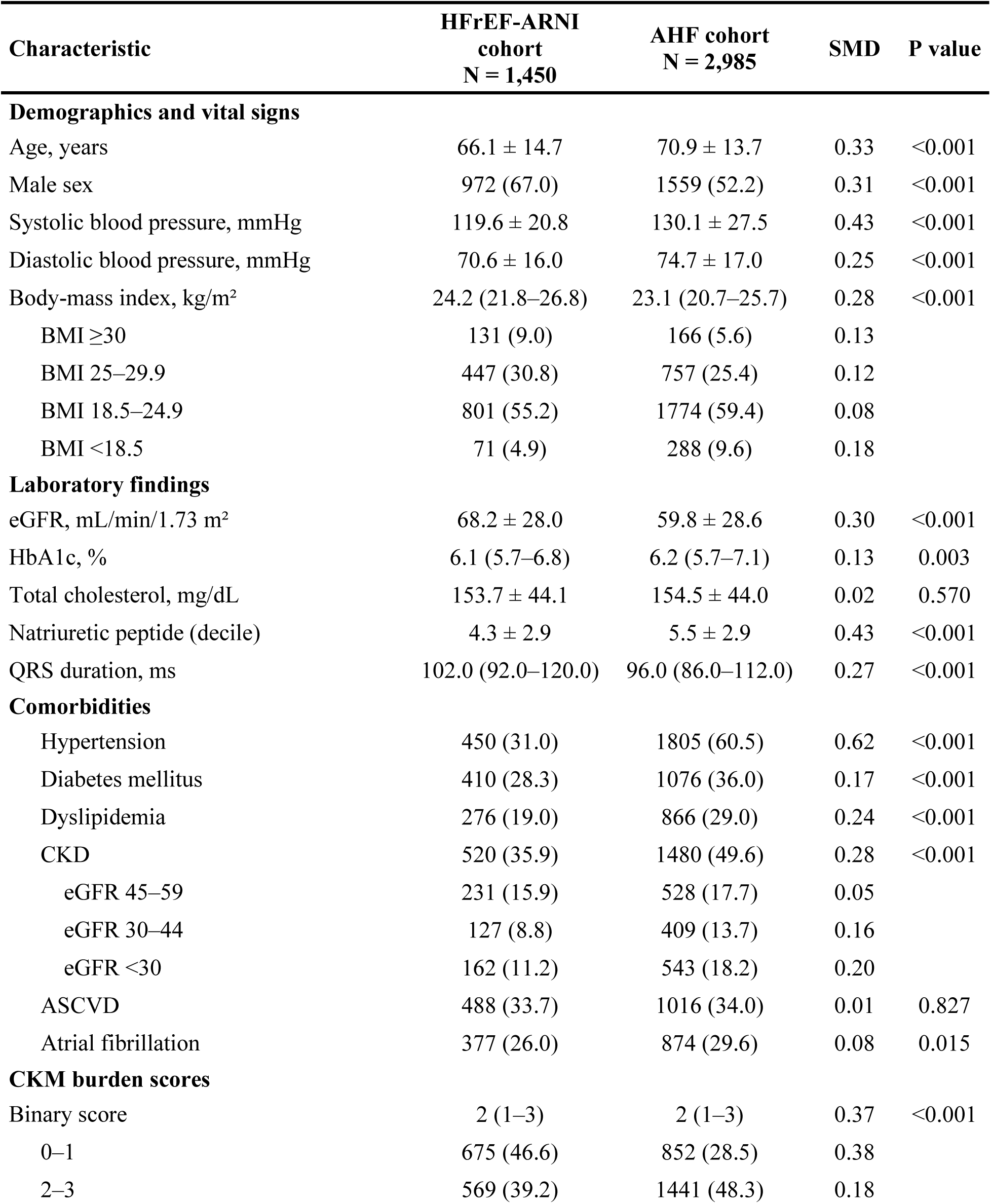
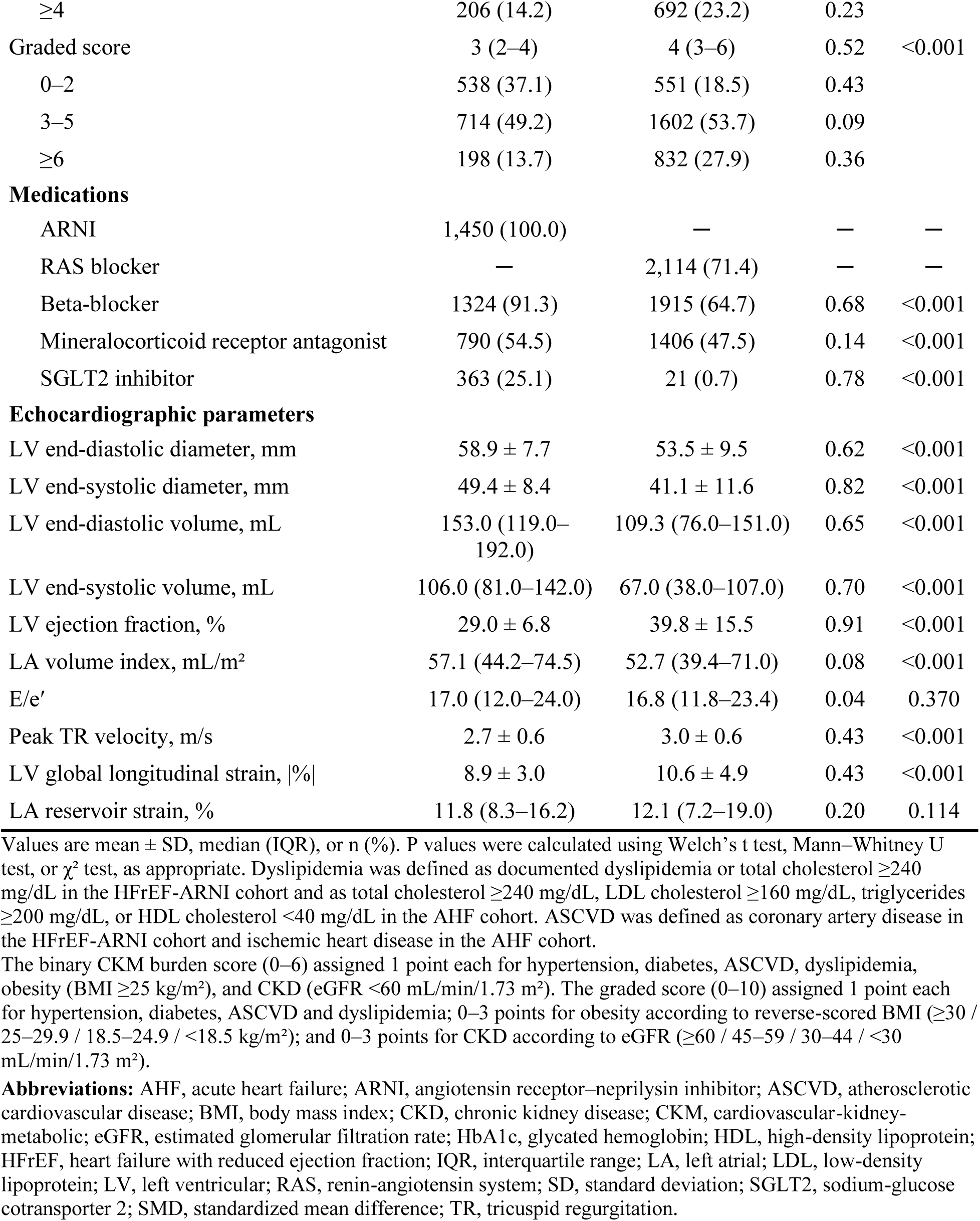
Baseline characteristics of the two cohorts.

Over a median follow-up of 31.5 months in the HFrEF-ARNI cohort and 51.1 months in the AHF cohort, the composite outcome occurred in 325 (22.9%) and 1,576 (52.8%), all-cause death in 128 (9.0%) and 1,244 (41.7%), and HF hospitalization in 245 (17.3%) and 608 patients (20.4%), respectively.

### CKM Burden and Clinical Outcomes

The binary CKM burden score reflected the number of coexisting CKM conditions, whereas the graded score incorporated CKD severity and reverse-scored BMI categories, reflecting their respective associations with adverse outcomes in both cohorts (**Supplemental Figure 1**). The two scoring approaches demonstrated different patterns of risk stratification. Three-year risk of the composite outcome increased progressively with increasing CKM burden assessed by the graded score in both cohorts (**Figure 1**). In contrast, the risk gradient was less pronounced with the binary score and tended to flatten at higher score levels. Similar patterns were observed for incidence rates per 100 person-years (**Supplemental Figure 2**), and for the individual components of the composite outcome (all-cause death and HF hospitalization; **Supplemental Figures 3 and 4**).

**Figure 1.**
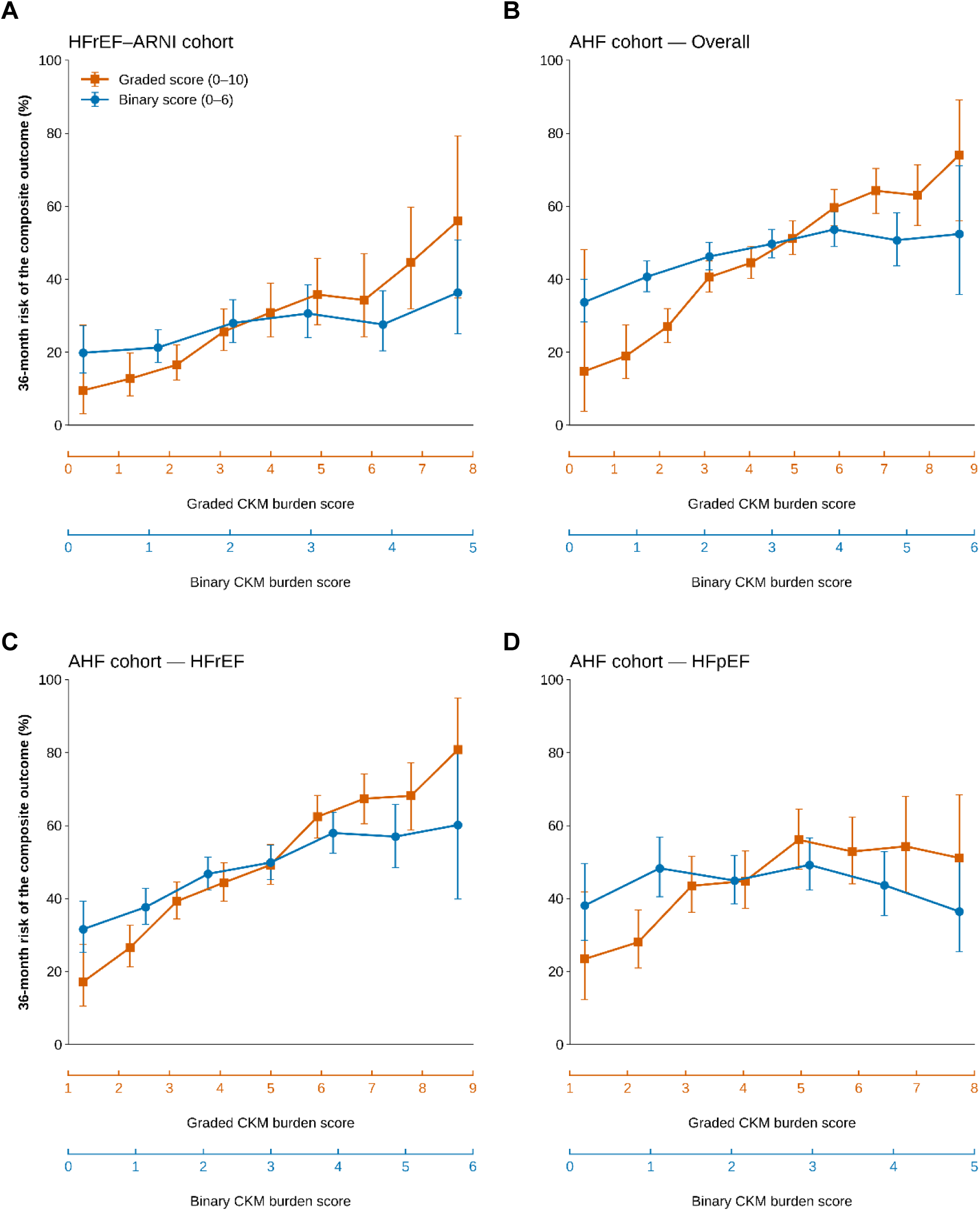
Three-Year Risk of the Composite Outcome across CKM Burden Scores. (A) HFrEF–ARNI cohort (n = 1,418); (B) AHF cohort – Overall (n = 2,984); (C) AHF cohort– HFrEF (n = 2,067); and (D) AHF cohort–HFpEF (n = 917). Unadjusted Kaplan–Meier estimates of the 36-month probability of the composite outcome of all-cause death or HF hospitalization are shown with 95% confidence intervals. Separate x-axis scales are used for the graded (orange) and binary (blue) CKM burden scores, reflecting their different score ranges. Score levels with fewer than 15 patients are not shown. Abbreviations: AHF, acute heart failure; ARNI, angiotensin receptor-neprilysin inhibitor; CKM, cardio-kidney-metabolic; HF, heart failure; HFpEF, heart failure with preserved ejection fraction; HFrEF, heart failure with reduced ejection fraction.

After multivariable adjustment, both scores were associated with the composite outcome in the HFrEF-ARNI cohort (graded: HR 1.11, 95% CI 1.04–1.19; binary: HR 1.10, 95% CI 1.02– 1.18) and AHF cohort (graded: HR 1.13, 95% CI 1.09–1.16; binary: HR 1.05, 95% CI 1.01– 1.09), with HRs expressed per 1-point increase in each score (**Table 2**, **Figure 2**). The graded score showed more consistent associations with all-cause death and HF hospitalization, whereas the binary score was not significantly associated with all-cause death in the HFrEF-ARNI cohort or HF hospitalization in the overall AHF cohort. Notably, in the HFpEF subgroup of the AHF cohort, the binary score was not significantly associated with any of the outcomes, whereas the graded score retained significant associations with the composite outcome and all-cause death, although these associations were weaker than those observed in the HFrEF subgroup (**Figure 2**).

**Figure 2.**
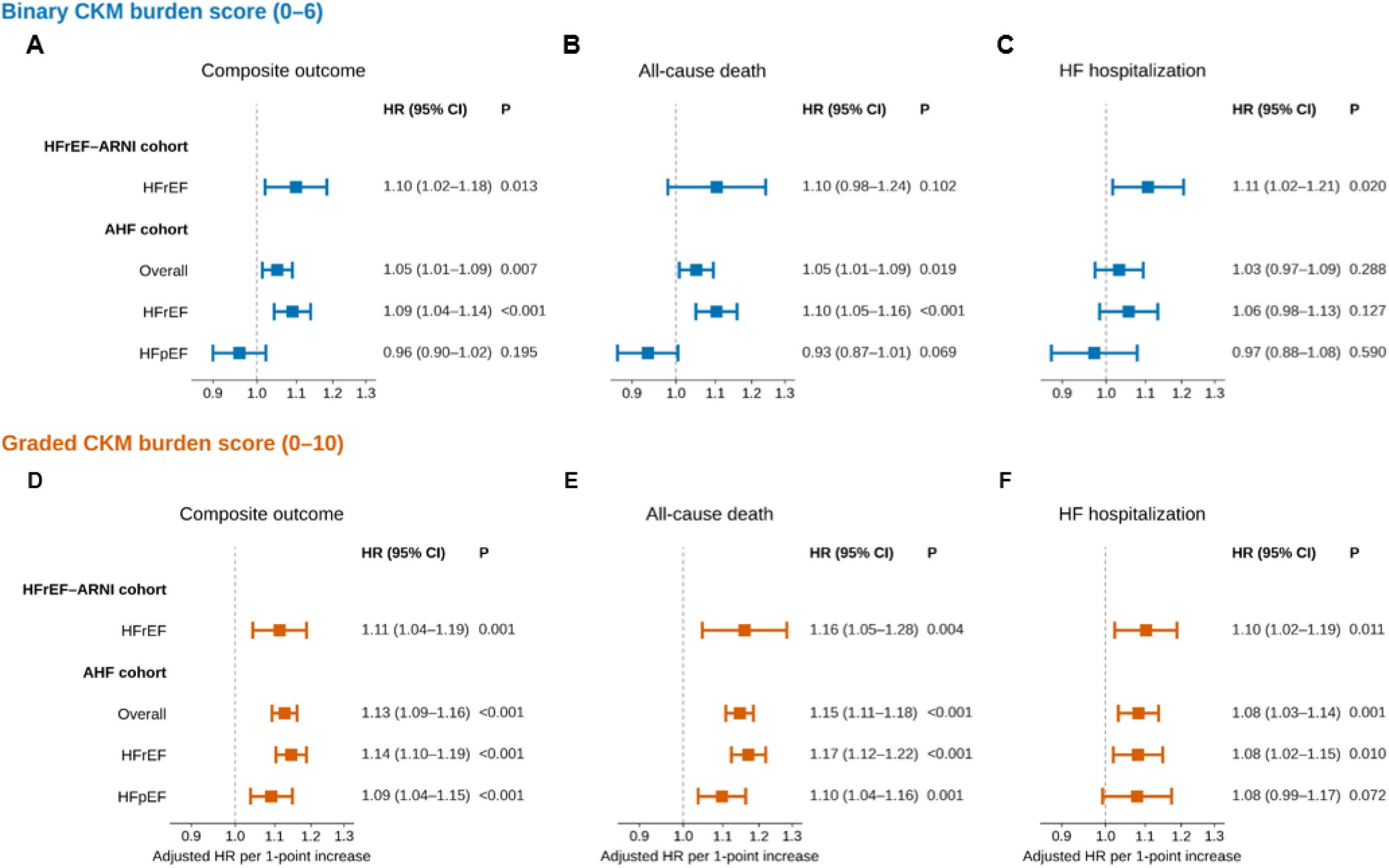
Association of CKM Burden Scores with the Clinical Outcomes. Panels A–C show the binary CKM burden score and panels D–F the graded CKM burden score, for the composite outcome (A, D), all-cause death (B, E), and HF hospitalization (C, F). Adjusted hazard ratios per 1-point increment in each CKM burden score are shown by cohort and HF category. The x-axis is on a logarithmic scale. Models were adjusted for age, sex, LVGLS, LASr, and natriuretic peptide level (decile). Abbreviations: CI, confidence interval; HR, hazard ratio; LASr, left atrial reservoir strain; LVGLS, left ventricular global longitudinal strain; others as in Figure 1.

**Table 2.**
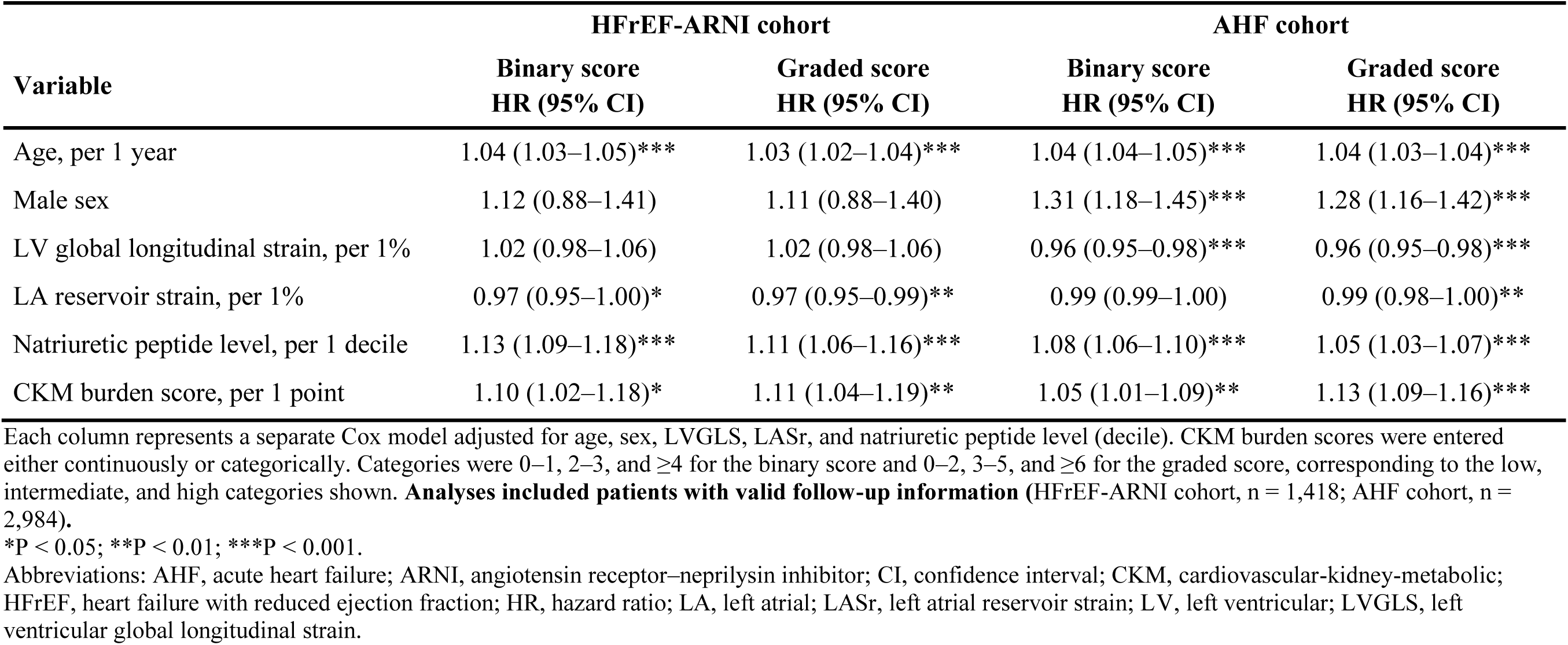
Multivariable Cox regression analysis for the primary composite outcome.

In categorical analyses, the graded score showed a more pronounced risk gradient, with adjusted HRs for the highest versus lowest category of 1.72 (95% CI 1.20–2.46) and 2.08 (95% CI 1.72–2.51) in the HFrEF-ARNI and AHF cohorts, respectively, compared with 1.36 (95% CI 1.00–1.85) and 1.19 (95% CI 1.03–1.37) for the binary score (**Figure 3, Supplemental Table 3**). Overall associations across categories were significant for the graded score in both cohorts (P = 0.011 and P <0.001), but not for the binary score (P = 0.057 and P = 0.051). Results for individual components of composite outcome are presented in **Supplemental Figures 5**.

**Figure 3.**
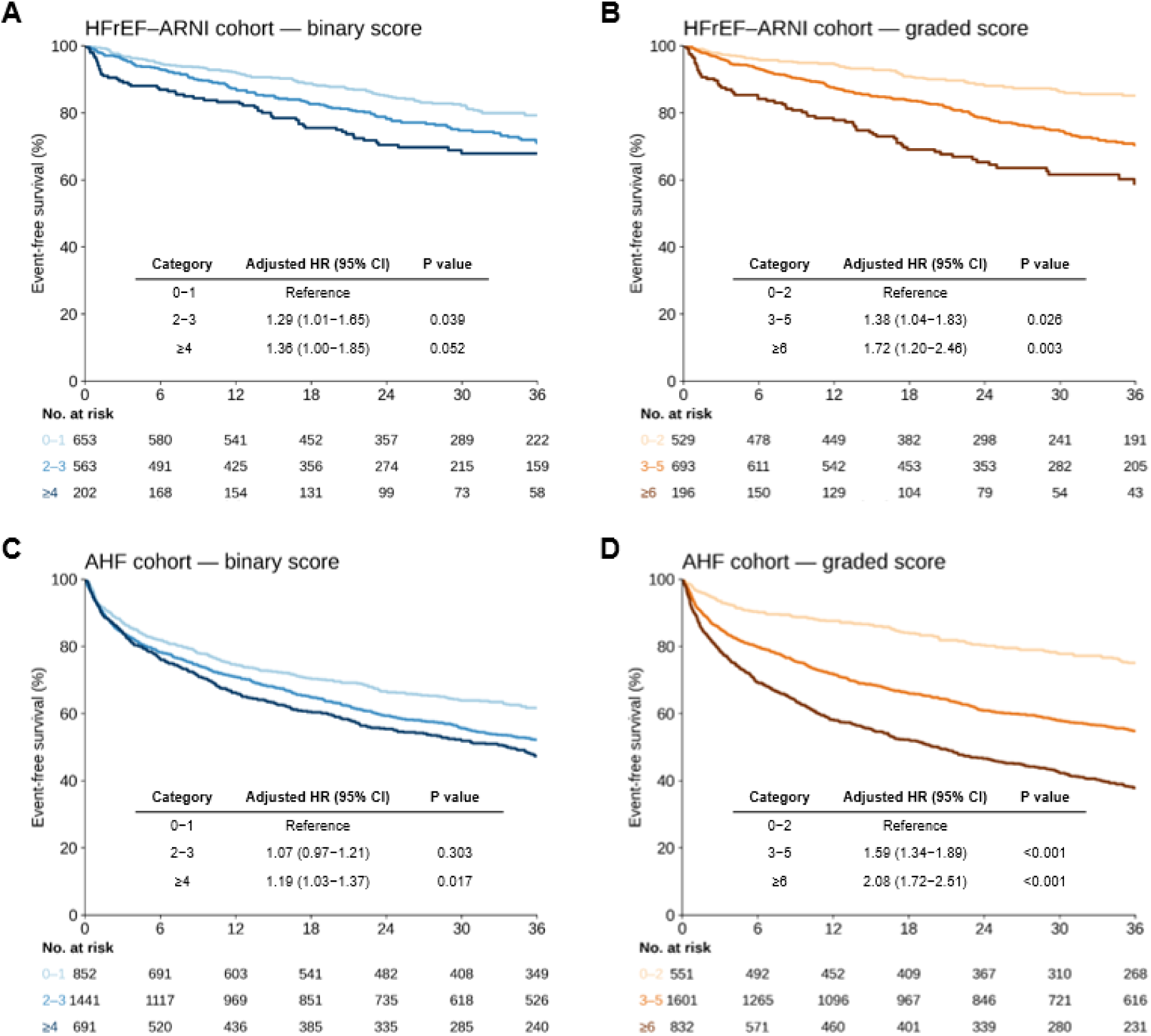
Event-free Survival of Composite Outcome by CKM Burden Score Categories. (A–B) HFrEF–ARNI cohort; (C–D) AHF cohort. Unadjusted Kaplan–Meier event-free survival is shown according to binary (A, C) and graded (B, D) CKM burden score categories. P values are from Cox models adjusted for age, sex, LVGLS, LASr, and natriuretic peptide level (decile). Score categories were 0–1, 2–3, and ≥4 for the binary score and 0–2, 3–5, and ≥6 for the graded score. Abbreviations as in Figure 1 and Figure 2.

**Figure 4.**
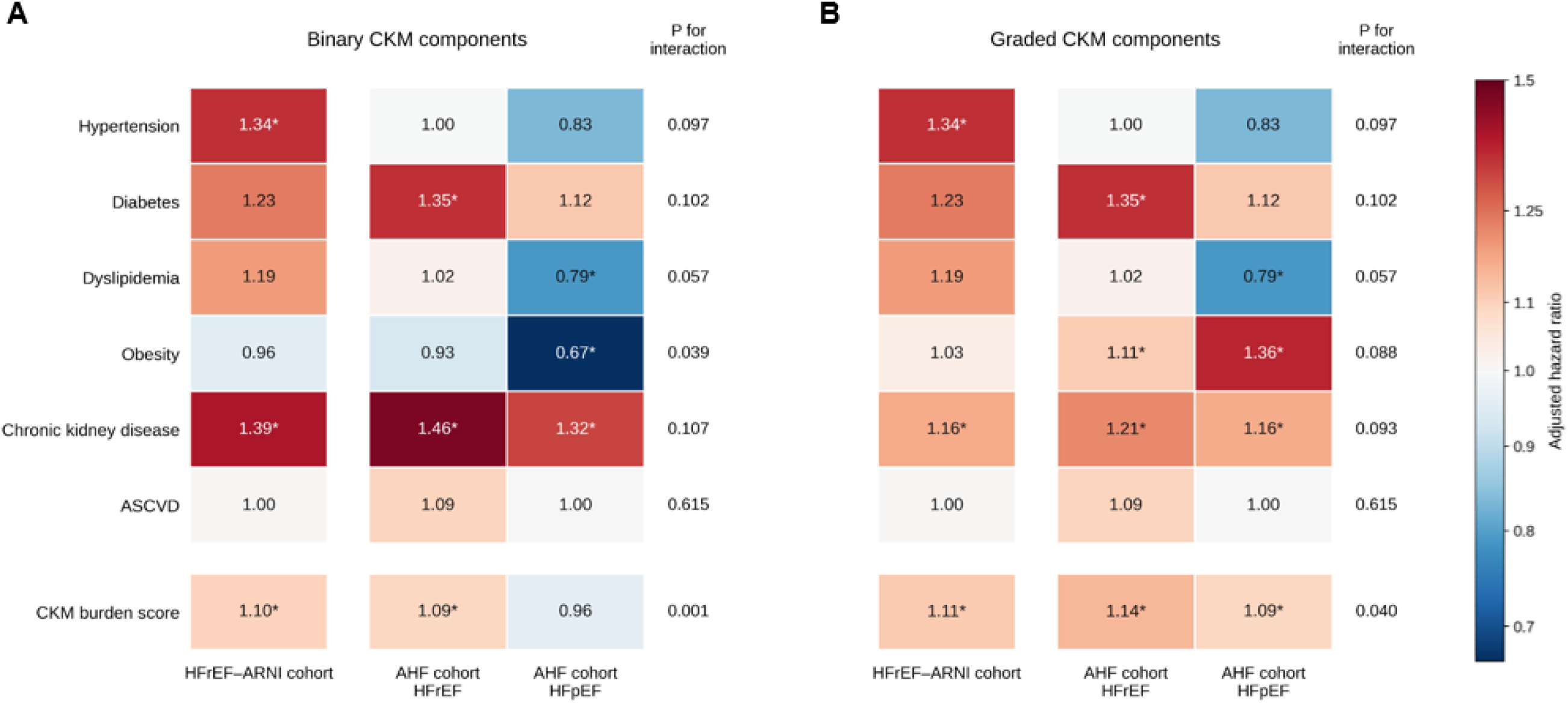
Associations of CKM Burden with Composite Outcome by HF Phenotype. (A) binary CKM components; (B) graded CKM components. Cells show adjusted hazard ratios for the composite outcome from separate Cox models for each CKM component, adjusted for age, sex, LVGLS, LASr, and natriuretic peptide level (decile). In panel A, hazard ratios represent the presence versus absence of each CKM component. In panel B, hazard ratios for CKD and obesity are expressed per 1-point increment in their respective graded scores (0–3) and are not directly comparable with the corresponding binary estimates. The remaining components were coded identically in both panels. The bottom row shows hazard ratios per 1-point increment in the overall CKM burden score. Columns are HFrEF–ARNI cohort and AHF cohort. P for interaction assesses whether the association differs between HFrEF and HFpEF within the AHF cohort. *P <0.05 within each subgroup. Abbreviations: ASCVD, atherosclerotic cardiovascular disease; CKD, chronic kidney disease; others as in Figure 1 and Figure 2.

In competing-risk analyses accounting for death before HF hospitalization, the subdistribution HR for the graded score was 1.04 (95% CI 1.00–1.10) in the AHF cohort, compared with a cause-specific HR of 1.08 (95% CI 1.03–1.14), in the presence of competing mortality (**Supplemental Figure 6, Supplemental Table 4**). Corresponding estimates were similar in the HFrEF-ARNI cohort (1.09 and 1.10, respectively).

### Prognostic Performance of CKM Burden Scores

The graded score showed higher C-indices than the binary score across outcomes and study populations (**Table 3 and Supplemental Table 5**). For the composite outcome, the C-index increased from 0.583 to 0.632 in the HFrEF-ARNI cohort (ΔC = 0.048, 95% CI 0.022–0.074) and from 0.541 to 0.609 in the AHF cohort (ΔC = 0.068, 95% CI 0.057–0.081). The largest improvement was observed in the HFpEF subgroup of the AHF cohort (ΔC = 0.085, 95% CI 0.062–0.108). The graded score also provided greater incremental model fit than the binary score. In the HFrEF-ARNI cohort, the likelihood-ratio χ² improvement was 10.3 for the graded score versus 6.1 for the binary score (P = 0.001 and P = 0.014, respectively). The corresponding values in the AHF cohort were 60.8 and 7.2 (P <0.001 and P = 0.007, respectively). Notably, in the HFpEF subgroup of the AHF cohort, only the graded score significantly improved model fit beyond the baseline model (Δχ² = 11.3, P <0.001; binary: Δχ² = 1.7, P = 0.195).

**Table 3.**
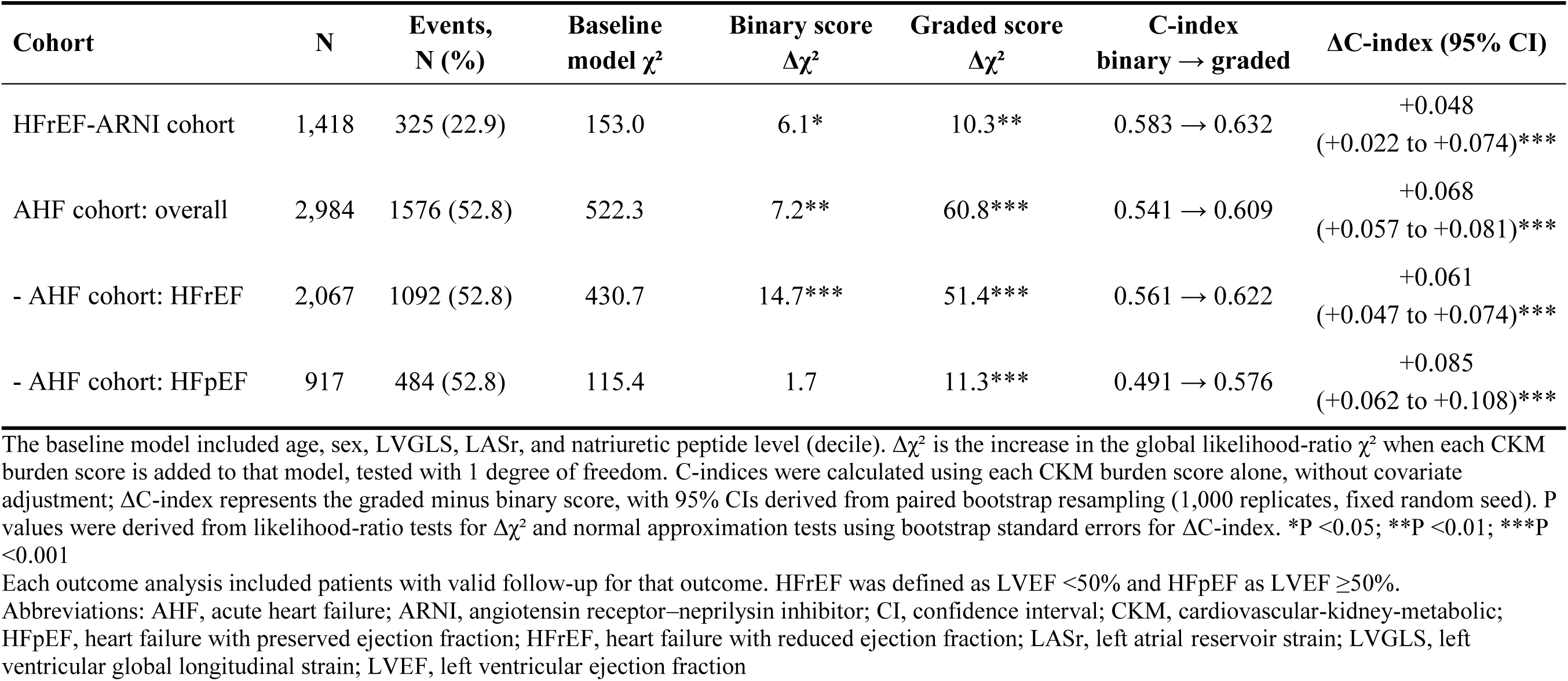
Incremental prognostic value of binary and graded CKM burden scores for the composite outcome.

| Cohort | N | Events, N (%) | Baseline model χ² | Binary score Δχ² | Graded score Δχ² | C-index binary → graded | ΔC-index (95% CI) |
|---|---|---|---|---|---|---|---|
| HFrEF-ARNI cohort | 1,418 | 325 (22.9) | 153.0 | 6.1* | 10.3** | 0.583 → 0.632 | +0.048 (+0.022 to +0.074)*** |
| AHF cohort: overall | 2,984 | 1576 (52.8) | 522.3 | 7.2** | 60.8*** | 0.541 → 0.609 | +0.068 (+0.057 to +0.081)*** |
| - AHF cohort: HFrEF | 2,067 | 1092 (52.8) | 430.7 | 14.7*** | 51.4*** | 0.561 → 0.622 | +0.061 (+0.047 to +0.074)*** |
| - AHF cohort: HFpEF | 917 | 484 (52.8) | 115.4 | 1.7 | 11.3*** | 0.491 → 0.576 | +0.085 (+0.062 to +0.108)*** |

The baseline model included age, sex, LVGLS, LASr, and natriuretic peptide level (decile). Δχ² is the increase in the global likelihood-ratio χ² when each CKM burden score is added to that model, tested with 1 degree of freedom. C-indices were calculated using each CKM burden score alone, without covariate adjustment; ΔC-index represents the graded minus binary score, with 95% CIs derived from paired bootstrap resampling (1,000 replicates, fixed random seed). P values were derived from likelihood-ratio tests for Δχ² and normal approximation tests using bootstrap standard errors for ΔC-index. \*P <0.05; \*\*P <0.01; \*\*\*P <0.001
Each outcome analysis included patients with valid follow-up for that outcome. HFrEF was defined as LVEF <50% and HFpEF as LVEF ≥50%.
Abbreviations: AHF, acute heart failure; ARNI, angiotensin receptor–neprilysin inhibitor; CI, confidence interval; CKM, cardiovascular-kidney-metabolic; HFpEF, heart failure with preserved ejection fraction; HFrEF, heart failure with reduced ejection fraction; LASr, left atrial reservoir strain; LVGLS, left ventricular global longitudinal strain; LVEF, left ventricular ejection fraction

### Prognostic Associations of Individual CKM Components Across HF Phenotypes

Given the significant interactions between CKM burden scores and HF phenotype (P-for-interaction=0.001 for the binary score and 0.040 for the graded score), we further examined the associations of individual CKM components with the composite outcome across cohorts and HF phenotypes (**Figure 4**). Among individual CKM components, CKD was consistently associated with higher risk of the composite outcome across cohorts and HF phenotypes using both binary and graded definitions. Hypertension was associated with higher risk in the HFrEF-ARNI cohort but not in either HF phenotype within the AHF cohort, whereas atherosclerotic cardiovascular disease showed no significant association across groups. In the HFpEF subgroup of the AHF cohort, dyslipidemia (HR 0.79, 95% CI 0.64–0.97) and binary obesity (BMI ≥25 kg/m²; HR 0.67, 95% CI 0.54–0.82) were associated with lower risk. In contrast, higher scores for the reverse-scored BMI component, indicating lower BMI, were associated with increased risk in the HFrEF (HR 1.11, 95% CI 1.01–1.22) and HFpEF (HR 1.36, 95% CI 1.19–1.56) subgroups of the AHF cohort.

### Network Analysis of CKM Components

Mixed graphical models showed conditional associations among CKM components, echocardiographic parameters, and clinical covariates (**Figure 5**). Across the four groups, connections were more evident within the CKM and echocardiographic domains than between them. Among CKM components, CKD showed the greatest prognostic contribution in the HFrEF-ARNI cohort, the overall AHF cohort, and the HFrEF subgroup of the AHF cohort, whereas obesity was most prominent in the HFpEF subgroup of the AHF cohort, mirroring the component-level associations shown in **Figure 4**.

**Figure 5.**
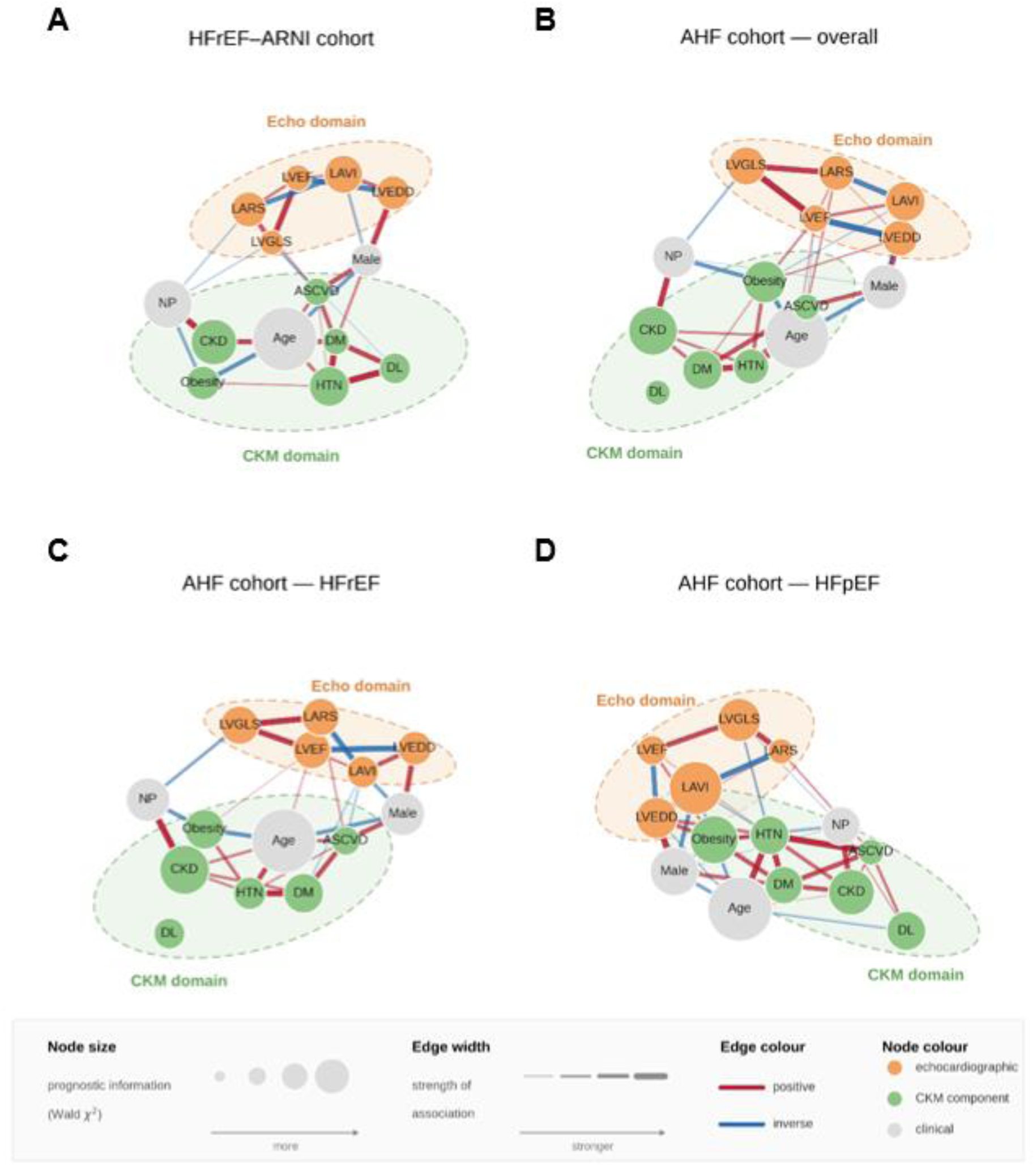
Network of Conditional Associations among CKM Components and Echocardiographic Parameters. (A) HFrEF–ARNI cohort; (B) AHF cohort – Overall; (C) AHF cohort–HFrEF; and (D) AHF cohort–HFpEF. Each panel shows a mixed graphical model estimated separately within each group using node-wise L1-regularized regression. Edges represent conditional associations retained using the AND rule and an edge-weight threshold of ≥0.10, with edge width indicating association strength and edge color indicating direction (red, positive; blue, inverse). Node area reflects the strength of the prognostic association from a multivariable Cox model for the composite outcome. Nodes are colored by domain: echocardiographic variables (orange), CKM components (green), and clinical variables (gray). Dashed ellipses indicate the echocardiographic and CKM domains. Node positions are not comparable across panels; variables without retained edges are displayed below the network. Abbreviations: DL, dyslipidemia; DM, diabetes mellitus; HTN, hypertension; LAVI, left atrial volume index; LVEDD, left ventricular end-diastolic dimension; LVEF, left ventricular ejection fraction; NP, natriuretic peptide; others as in Figure 1 and Figure 2.

## Discussion

In two independent HF cohorts, an HF-adapted graded CKM burden score incorporating CKD severity and reverse-scored BMI categories demonstrated more consistent prognostic associations than a binary CKM burden score based on the same six components. The graded score showed better discrimination when evaluated alone and greater incremental model fit beyond clinical and echocardiographic variables. The distinction was particularly evident in HFpEF, where only the graded score was significantly associated with the composite outcome. These findings support the prognostic relevance of CKM burden in established HF and suggest that its assessment should consider not only the presence of CKM conditions but also disease severity and HF-relevant prognostic characteristics.

### CKM Burden Assessment in Established HF

The CKM framework classifies established HF in the presence of metabolic risk factors or CKD as stage 4 CKM syndrome, while its quantitative risk assessment recommendations primarily focus on predicting cardiovascular disease development, including HF.^1^ The metabolic and kidney abnormalities that contribute to HF development remain clinically relevant after its onset, providing a rationale for evaluating their cumulative prognostic implications in established HF. The high prevalence of CKM conditions in our cohorts further underscores the relevance of this assessment.

Previous studies have demonstrated the prognostic importance of CKM multimorbidity by counting coexisting conditions using binary definitions.^3–8^ However, this approach may not adequately capture differences in disease severity or the prognostic characteristics of individual components in established HF. Indeed, conventional cardiovascular risk factors may have different prognostic implications before and after HF development.^18–20^ For example, more advanced kidney dysfunction and lower BMI have been associated with increased risk in patients with established HF, highlighting the importance of disease severity and the distinct prognostic characteristics of cardiometabolic risk factors in this population.^20, 21^ Therefore, while the CKM framework remains relevant after HF is established, its conventional assessment based on the presence of individual components may require refinement to better characterize their prognostic implications.

### Severity-Based and HF-Adapted CKM Assessment

Kidney dysfunction is a major prognostic factor in HF, and its severity provides information beyond the presence or absence of CKD.^17, 21^ Similarly, an inverse association between BMI and mortality has been reported in patients with established HF, commonly described as the obesity paradox.^20^ Consistent with these observations, greater kidney dysfunction and lower BMI were associated with higher risk of adverse outcomes in both cohorts. These findings informed our incorporation of eGFR-based CKD categories and reverse-scored BMI categories into the graded CKM burden score.

Compared with the binary score, the graded score demonstrated clearer risk gradients, more consistent associations with clinical outcomes, and better prognostic discrimination when evaluated alone. It also provided greater incremental model fit beyond clinical and echocardiographic variables. Importantly, the two modifications represent different aspects of prognostic assessment: CKD grading captures increasing disease severity, whereas reverse-scored BMI captures the direction of an observed risk association. Thus, the graded score should be interpreted as an HF-adapted measure of prognostic burden rather than simply a measure of increasing cardiometabolic disease severity.

Exploratory network analysis demonstrated that conditional associations were more frequent within than between CKM and echocardiographic domains. These findings suggest that CKM components and echocardiographic parameters may reflect partly distinct aspects of clinical status. Together, these findings suggest that an HF-adapted CKM burden assessment incorporating disease severity and HF-relevant prognostic characteristics may complement conventional clinical and echocardiographic evaluation in patients with established HF.

### CKM Burden Across HF Phenotypes

The distinction between the scoring approaches was particularly evident in HFpEF. Consistent with previous registry studies reporting weaker prognostic associations of binary-defined CKM multimorbidity in HFpEF than in HFrEF,^3, 8^ the binary score was associated with the composite outcome in the HFrEF subgroup of the AHF cohort but not in HFpEF. In contrast, the graded score retained significant associations in both phenotypes and improved model fit in HFpEF. Nevertheless, significant interactions with HF phenotype were observed for both scores, indicating that the graded approach did not completely eliminate phenotype-dependent differences in their prognostic associations.

Component-level analyses may provide insight into these phenotype-specific differences. Within the AHF cohort, diabetes was more prevalent and was associated with adverse outcomes in HFrEF, whereas hypertension and obesity were more prevalent in HFpEF but did not demonstrate corresponding positive associations with clinical risk. Dyslipidemia was also associated with lower risk in HFpEF. These patterns may weaken the prognostic association of a binary score that assigns positive points to the presence of these conditions, especially in HFpEF. Notably, the inverse association of binary-defined obesity, consistent with the obesity paradox in established HF,^21^ was more pronounced in HFpEF, with a significant interaction by HF phenotype. Incorporating CKD severity and reversing the direction of BMI scoring may therefore partly explain the retained prognostic association of the graded score in HFpEF. Network analysis also identified the BMI-related obesity component as the CKM component with the greatest prognostic contribution in HFpEF, compared with CKD in HFrEF, further highlighting the potential importance of BMI characterization in phenotype-specific prognostic assessment.

These findings suggest that simply counting CKM conditions may not adequately capture their prognostic implications in established HF. CKM components may have different prognostic characteristics according to HF phenotype, and their associations with outcomes may differ from their conventional roles as risk factors for HF development. Accordingly, the assessment of CKM burden across the HF spectrum may benefit from incorporating component severity and HF-relevant prognostic characteristics rather than relying solely on conventional binary definitions.

### Limitations

Several limitations should be acknowledged. First, the retrospective observational design precludes causal inference, and residual confounding cannot be excluded. Second, the graded score was developed and evaluated in the same cohorts; therefore, its prognostic performance requires confirmation in independent populations. Third, component assessment was constrained by available data. The atherosclerotic cardiovascular disease component was restricted to coronary disease, and dyslipidemia definitions differed between cohorts. Severity grading was not incorporated for the remaining CKM components, and whether further characterization would improve performance remains uncertain. Fourth, CKD grading and BMI reverse-scoring increased the relative weighting of these components. Their separate contributions to the improvement in prognostic performance could not be determined, and alternative scoring approaches were not comprehensively evaluated. Fifth, medication information was limited to baseline use, and treatment patterns differed between cohorts, partly reflecting their enrollment periods. Finally, both cohorts predominantly comprised Asian patients, and the applicability of the score to other populations requires further evaluation.

## Conclusion

An HF-adapted graded CKM burden score, incorporating CKD severity and reverse-scored BMI categories, demonstrated more consistent prognostic associations and better risk discrimination than the binary score across two HF cohorts. These differences were particularly evident in HFpEF, where the binary score showed limited prognostic value. Our findings support the prognostic value of CKM burden in established HF and highlight the importance of considering disease severity and HF-relevant prognostic characteristics in its assessment.

## Clinical Perspectives

### Competency in Medical Knowledge

CKM burden may inform prognostic assessment across the HF spectrum. An HF-adapted graded CKM burden score incorporating CKD severity and reverse-scored BMI categories demonstrated more consistent prognostic associations and better risk discrimination than a binary score based on the presence of CKM conditions. This distinction was particularly relevant in HFpEF, where the binary score showed limited prognostic value.

### Translational Outlook

More detailed information on CKM component severity and medication use may help refine HF-adapted CKM burden scoring. Prospective studies are needed to evaluate its clinical utility for risk stratification and patient management in HF.

## Data Availability

The data underlying this study are available from the corresponding author upon reasonable request, subject to approval and applicable ethical and privacy restrictions.

## Abbreviations

AHF: acute heart failure
ARNI: angiotensin receptor–neprilysin inhibitor
ASCVD: atherosclerotic cardiovascular disease
BMI: body mass index
CI: confidence interval
CKD: chronic kidney disease
CKM: cardiovascular-kidney-metabolic
DL: dyslipidemia
DM: diabetes mellitus
HF: heart failure
HfpEF: heart failure with preserved ejection fraction
HfrEF: heart failure with reduced ejection fraction
HR: hazard ratio
HTN: hypertension.

## Supplemental Materials

### Supplemental Methods

#### Supplemental Method 1. CKM Burden Score Construction and Categorization

A binary cardiovascular-kidney-metabolic (CKM) burden score assigned 1 point each for hypertension, diabetes, atherosclerotic cardiovascular disease (ASCVD), dyslipidemia, obesity, and chronic kidney disease (CKD), yielding a total score of 0–6. Obesity was defined as body mass index (BMI) ≥25 kg/m² and CKD as estimated glomerular filtration rate (eGFR) <60 mL/min/1.73 m². For the heart failure (HF)-adapted graded CKM burden score, hypertension, diabetes, ASCVD, and dyslipidemia retained binary scoring, whereas CKD and obesity were each assigned 0–3 points, yielding a total score of 0–10. CKD was scored according to eGFR (≥60, 45–59, 30–44, and <30 mL/min/1.73 m²: 0, 1, 2, and 3 points, respectively), and BMI was reverse-scored for the obesity component (≥30, 25–29.9, 18.5–24.9, and <18.5 kg/m²: 0, 1, 2, and 3 points, respectively), consistent with the obesity paradox observed in heart failure.^1^

Component definitions were based on available data in each cohort. The ASCVD component was based on coronary artery disease in the HF with reduced ejection fraction (HFrEF) cohort and ischemic heart disease in the acute HF (AHF) cohort. Dyslipidemia was defined as a recorded diagnosis or total cholesterol ≥240 mg/dL in the HFrEF-ARNI cohort. In the AHF cohort, where diagnostic information on dyslipidemia was unavailable, it was defined as total cholesterol ≥240 mg/dL, low-density lipoprotein cholesterol ≥160 mg/dL, triglycerides ≥200 mg/dL, or high-density lipoprotein cholesterol <40 mg/dL.

Candidate category boundaries were evaluated based on observed risk gradients across integer score levels in both cohorts, category sizes, and model fit. Each category was required to include at least 50 patients in both cohorts. The selected boundaries showed comparable model fit to the best-performing alternatives (ΔAIC = 1.8 for the binary score and 1.5 for the graded score), with greater emphasis placed on risk gradients and category distributions. The final categories were 0–1, 2–3, and ≥4 for the binary score and 0–2, 3–5, and ≥6 for the graded score, with identical boundaries applied in both cohorts to facilitate clinical interpretation.

#### Supplemental Method 2. Details of the Prognostic and Network Analyses

Time-to-event associations were assessed using Cox proportional hazards models adjusted for age, sex, left ventricular global longitudinal strain (LVGLS), left atrial reservoir strain (LASr), and natriuretic peptide (NP) decile levels. B-type natriuretic peptide (BNP) and N-terminal pro-BNP (NT-proBNP) levels were categorized into a common NP decile scale using previously published cutoffs (**Supplemental Table 1**).^2^ Analyses were performed in each cohort and separately by HF phenotype within the AHF cohort. Differences across CKM burden score categories were assessed using likelihood-ratio tests with 2 degrees of freedom in adjusted Cox models.

Proportional hazards assumptions were examined using scaled Schoenfeld residuals. Unadjusted absolute risks at 36 months were estimated using the Kaplan–Meier method with 95% confidence intervals, and incidence rates were calculated per 100 person-years with exact Poisson 95% confidence intervals. For HF hospitalization, cumulative incidence was estimated using the Aalen–Johansen method, with death as a competing event. Subdistribution hazard ratios were estimated using Fine–Gray models, retaining subjects who experienced the competing event in the risk set through inverse-probability-of-censoring weighting. Variance was estimated using a robust sandwich variance estimator.

Median follow-up was estimated using the reverse Kaplan–Meier method. Discrimination was assessed using Harrell’s C-index for each CKM burden score alone. Differences in C-index between the binary and graded scores were evaluated using paired bootstrap resampling (1,000 replicates), with percentile 95% confidence intervals and P values derived using a normal approximation based on bootstrap standard errors. Incremental prognostic value was assessed by adding each CKM burden score to the baseline model (age, sex, LVGLS, LASr, and NP decile) and comparing model fit using likelihood-ratio tests with 1 degree of freedom.

Mixed graphical models were estimated separately in the HFrEF-ARNI cohort, the overall AHF cohort, and the HFrEF and HF with preserved ejection fraction (HFpEF) subgroups of the AHF cohort. Each of the 14 variables was regressed on all others using L1-regularized linear regression for continuous variables and logistic regression for binary variables. The regularization parameter was selected using 5-fold cross-validation with a fixed random seed. An edge was retained only when both node-wise coefficients were nonzero and the symmetrized edge weight, defined as √(|βab| × |βba|), was ≥0.10. Node area was scaled with the square root of the Wald χ² statistic for each variable from a multivariable Cox model for the composite outcome containing all 14 variables, representing prognostic information independently of network position. Network analyses were restricted to patients with complete data for all 14 variables.

## Supplemental Tables

**Supplemental Table 1.** Cutoff values for deciles of natriuretic peptide levels.

| **Decile** | **BNP (pg/mL)** | **NT-proBNP (pg/mL)** |
|---|---|---|
| **1** | <160 | <579.9 |
| **2** | 160–<346 | 579.9–<1335 |
| **3** | 346–<540 | 1335–<2119 |
| **4** | 540–<739 | 2119–<3217 |
| **5** | 739–<980 | 3217–<4583.6 |
| **6** | 980–<1279 | 4583.6–<6274.8 |
| **7** | 1279–<1746 | 6274.8–<9214 |
| **8** | 1746–<2530 | 9214–<14293.2 |
| **9** | 2530–<3629 | 14293.2–<27924.9 |
| **10** | ≥3629 | ≥27924.9 |

Natriuretic peptide level (decile) cutoffs were adopted from previously published data.<sup>1</sup>
Abbreviations: BNP, B-type natriuretic peptide; NT-proBNP, N-terminal pro-B-type natriuretic peptide.

**Supplemental Table 2.** Baseline Characteristics of the AHF cohort According to HF phenotype.

| Characteristic | Overall<br>N = 2,985 | HFrEF<br>N = 2,067 | HFpEF<br>N = 918 | SMD | P value |
|---|---|---|---|---|---|
| **Demographics and vital signs** |  |  |  |  |  |
| Age, years | 70.9 ± 13.7 | 69.6 ± 13.9 | 73.7 ± 12.6 | 0.31 | <0.001 |
| Male sex | 1559 (52.2) | 1198 (58.0) | 361 (39.3) | 0.38 | <0.001 |
| Systolic blood pressure, mmHg | 130.1 ± 27.5 | 128.8 ± 27.0 | 133.1 ± 28.5 | 0.16 | <0.001 |
| Diastolic blood pressure, mmHg | 74.7 ± 17.0 | 75.3 ± 17.0 | 73.4 ± 16.9 | 0.11 | 0.005 |
| Body mass index, kg/m² | 23.1 (20.7–25.7) | 22.9 (20.5–25.5) | 23.5 (21.1–26.2) | 0.14 | <0.001 |
| BMI ≥30 | 166 (5.6) | 102 (4.9) | 64 (7.0) | 0.09 |  |
| BMI 25–29.9 | 757 (25.4) | 497 (24.0) | 260 (28.3) | 0.10 |  |
| BMI 18.5–24.9 | 1774 (59.4) | 1259 (60.9) | 515 (56.1) | 0.10 |  |
| BMI <18.5 | 288 (9.6) | 209 (10.1) | 79 (8.6) | 0.05 |  |
| **Laboratory findings** |  |  |  |  |  |
| eGFR, mL/min/1.73 m² | 59.8 ± 28.6 | 59.9 ± 28.7 | 59.5 ± 28.4 | 0.02 | 0.695 |
| HbA1c, % | 6.2 (5.7–7.1) | 6.3 (5.7–7.2) | 6.1 (5.7–6.8) | 0.14 | 0.002 |
| Total cholesterol, mg/dL | 154.5 ± 44.0 | 154.7 ± 43.7 | 154.0 ± 44.7 | 0.02 | 0.705 |
| Natriuretic peptide (decile) | 5.5 ± 2.9 | 6.0 ± 2.8 | 4.5 ± 2.7 | 0.54 | <0.001 |
| QRS duration, ms | 96.0 (86.0–112.0) | 98.0 (88.0–116.0) | 91.0 (82.0–102.0) | 0.33 | <0.001 |
| **Comorbidities** |  |  |  |  |  |
| Hypertension | 1805 (60.5) | 1202 (58.2) | 603 (65.7) | 0.16 | <0.001 |
| Diabetes mellitus | 1076 (36.0) | 776 (37.5) | 300 (32.7) | 0.10 | 0.012 |
| Dyslipidemia | 866 (29.0) | 617 (29.9) | 249 (27.1) | 0.06 | 0.141 |
| CKD | 1480 (49.6) | 1022 (49.4) | 458 (49.9) | 0.01 | 0.852 |
| eGFR 45–59 | 528 (17.7) | 360 (17.4) | 168 (18.3) | 0.02 |  |
| eGFR 30–44 | 409 (13.7) | 289 (14.0) | 120 (13.1) | 0.03 |  |
| eGFR <30 | 543 (18.2) | 373 (18.0) | 170 (18.5) | 0.01 |  |
| ASCVD | 1016 (34.0) | 747 (36.1) | 269 (29.3) | 0.15 | <0.001 |
| Atrial fibrillation | 874 (29.6) | 551 (27.0) | 323 (35.3) | 0.18 | <0.001 |
| **CKM burden scores** |  |  |  |  |  |
| Binary score, median (IQR) | 2 (1–3) | 2 (1–3) | 2 (1–3) | 0.00 | 0.991 |
| 0–1 | 852 (28.5) | 592 (28.6) | 260 (28.3) | 0.01 |  |
| 2–3 | 1441 (48.3) | 998 (48.3) | 443 (48.3) | 0.00 |  |
| ≥4 | 692 (23.2) | 477 (23.1) | 215 (23.4) | 0.01 |  |
| Graded score, median (IQR) | 4 (3–6) | 4 (3–6) | 4 (3–6) | 0.09 | 0.026 |
| 0–2 | 551 (18.5) | 366 (17.7) | 185 (20.2) | 0.06 |  |
| 3–5 | 1602 (53.7) | 1102 (53.3) | 500 (54.5) | 0.02 |  |
| ≥6 | 832 (27.9) | 599 (29.0) | 233 (25.4) | 0.08 |  |
| **Medications** |  |  |  |  |  |
| RAS blocker | 2,114 (71.4) | 1,565 (76.4) | 549 (60.2) | 0.35 | <0.001 |
| Beta-blocker | 1915 (64.7) | 1395 (68.1) | 520 (57.0) | 0.23 | <0.001 |
| Mineralocorticoid receptor antagonist | 1406 (47.5) | 1055 (51.5) | 351 (38.5) | 0.26 | <0.001 |
| SGLT2 inhibitor | 21 (0.7) | 17 (0.8) | 4 (0.4) | 0.05 | 0.353 |
| **Echocardiographic parameters** |  |  |  |  |  |
| LV end-diastolic diameter, mm | 53.5 ± 9.5 | 56.3 ± 9.1 | 47.3 ± 7.1 | 1.11 | <0.001 |
| LV end-systolic diameter, mm | 41.1 ± 11.6 | 46.0 ± 10.3 | 31.0 ± 6.4 | 1.75 | <0.001 |
| LV end-diastolic volume, mL | 109.3 (76.0–151.0) | 126.3 (94.5–168.4) | 72.9 (56.4–95.0) | 1.20 | <0.001 |
| LV end-systolic volume, mL | 67.0 (38.0–107.0) | 86.0 (59.0–123.0) | 30.0 (21.5–40.1) | 1.62 | <0.001 |
| LV ejection fraction, % | 39.8 ± 15.5 | 31.4 ± 9.8 | 58.9 ± 5.8 | 3.42 | <0.001 |
| LA volume index, mL/m² | 52.7 (39.4–71.0) | 52.7 (40.3–69.7) | 52.5 (36.3–73.7) | 0.07 | 0.406 |
| E/e′ | 16.8 (11.8–23.4) | 17.7 (12.5–24.6) | 14.8 (11.0–20.8) | 0.28 | <0.001 |
| Peak tricuspid regurgitation velocity, m/s | 3.0 ± 0.6 | 3.0 ± 0.6 | 2.9 ± 0.6 | 0.09 | 0.090 |
| LV global longitudinal strain, \|%\| | 10.6 ± 4.9 | 8.7 ± 3.6 | 15.0 ± 4.5 | 1.56 | <0.001 |
| LA reservoir strain, % | 12.1 (7.2–19.0) | 10.5 (6.5–16.0) | 17.6 (10.1–26.0) | 0.68 | <0.001 |

Values are presented as mean ± SD, median (IQR), or n (%). P values compare HFrEF with HFpEF and were calculated using Welch's t test, the Mann–Whitney U test, or the χ² test, as appropriate; the overall column is descriptive and was not included in the comparison. HFrEF was defined as LVEF <50% and HFpEF as LVEF ≥50%. Dyslipidemia was defined as total cholesterol ≥240 mg/dL, LDL cholesterol ≥160 mg/dL, triglycerides ≥200 mg/dL, or HDL cholesterol <40 mg/dL in AHF cohort. ASCVD was defined as coronary or ischemic heart disease.
The binary CKM burden score (0–6) assigned 1 point each for hypertension, diabetes, ASCVD, dyslipidemia, obesity (BMI ≥25 kg/m²), and CKD (eGFR <60 mL/min/1.73 m²).
The graded score (0–10) assigned 1 point each for hypertension, diabetes, ASCVD, and dyslipidemia; 0–3 points for obesity according to reverse-scored BMI (≥30 / 25–29.9 / 18.5–24.9 / <18.5 kg/m²); and 0–3 points for CKD according to eGFR (≥60 / 45–59 / 30–44 / <30 mL/min/1.73 m²).
**Abbreviations:** AHF, acute heart failure; ASCVD, atherosclerotic cardiovascular disease; BMI, body mass index; CKD, chronic kidney disease; CKM, cardiovascular-kidney-metabolic; eGFR, estimated glomerular filtration rate; HbA1c, glycated hemoglobin; HDL, high-density lipoprotein; HF, heart failure; HFpEF, heart failure with preserved ejection fraction; HFrEF, heart failure with reduced ejection fraction; IQR, interquartile range; LA, left atrial; LDL, low-density lipoprotein; LV, left ventricular; LVEF, left ventricular ejection fraction; RAS, renin-angiotensin system; SD, standard deviation; SGLT2, sodium-glucose cotransporter 2; SMD, standardized mean difference.

**Supplemental Table 3.** Multivariable Associations and Prognostic Performance of Binary and Graded CKM Burden Score Categories.

| Outcome | Cohort | Binary CKM burden score (0–6) |  |  | Graded CKM burden score (0–10) |  |  |
|---|---|---|---|---|---|---|---|
|  |  | Category | Events<br>N (%) | Adjusted HR<br>(95% CI) | Category | Events<br>N (%) | Adjusted HR<br>(95% CI) |
| Composite outcome | HFrEF-ARNI cohort | 0–1 | 122 (18.7) | 1.00 (reference) | 0–2 | 75 (14.2) | 1.00 (reference) |
|  |  | 2–3 | 140 (24.9) | 1.29 (1.01–1.65)* | 3–5 | 180 (26.0) | 1.38 (1.04–1.83)* |
|  |  | ≥4 | 63 (31.2) | 1.36 (1.00–1.85) | ≥6 | 70 (35.7) | 1.72 (1.20–2.46)** |
|  | AHF cohort: overall | 0–1 | 369 (43.3) | 1.00 (reference) | 0–2 | 157 (28.5) | 1.00 (reference) |
|  |  | 2–3 | 786 (54.5) | 1.07 (0.94–1.21) | 3–5 | 843 (52.7) | 1.59 (1.34–1.89)*** |
|  |  | ≥4 | 421 (60.9) | 1.19 (1.03–1.37)* | ≥6 | 576 (69.2) | 2.08 (1.72–2.51)*** |
|  | - AHF cohort: HFrEF | 0–1 | 246 (41.6) | 1.00 (reference) | 0–2 | 97 (26.5) | 1.00 (reference) |
|  |  | 2–3 | 543 (54.4) | 1.12 (0.96–1.31) | 3–5 | 571 (51.8) | 1.59 (1.28–1.99)*** |
|  |  | ≥4 | 303 (63.5) | 1.36 (1.14–1.61)*** | ≥6 | 424 (70.8) | 2.23 (1.75–2.83)*** |
|  | - AHF cohort: HFpEF | 0–1 | 123 (47.3) | 1.00 (reference) | 0–2 | 60 (32.4) | 1.00 (reference) |
|  |  | 2–3 | 243 (54.9) | 0.95 (0.76–1.18) | 3–5 | 272 (54.5) | 1.67 (1.25–2.22)*** |
|  |  | ≥4 | 118 (55.1) | 0.83 (0.64–1.07) | ≥6 | 152 (65.2) | 1.80 (1.32–2.46)*** |
| All-cause death | HFrEF-ARNI cohort | 0–1 | 44 (6.7) | 1.00 (reference) | 0–2 | 21 (4.0) | 1.00 (reference) |
|  |  | 2–3 | 55 (9.8) | 1.28 (0.86–1.91) | 3–5 | 67 (9.7) | 1.40 (0.84–2.35) |
|  |  | ≥4 | 29 (14.3) | 1.37 (0.85–2.21) | ≥6 | 40 (20.4) | 2.21 (1.22–3.98)** |
|  | AHF cohort: overall | 0–1 | 283 (33.2) | 1.00 (reference) | 0–2 | 105 (19.1) | 1.00 (reference) |
|  |  | 2–3 | 608 (42.2) | 1.01 (0.87–1.16) | 3–5 | 639 (39.9) | 1.61 (1.31–1.99)*** |
|  |  | ≥4 | 353 (51.0) | 1.21 (1.03–1.42)* | ≥6 | 500 (60.1) | 2.23 (1.78–2.78)*** |
|  | - AHF cohort: HFrEF | 0–1 | 182 (30.7) | 1.00 (reference) | 0–2 | 64 (17.5) | 1.00 (reference) |
|  |  | 2–3 | 423 (42.4) | 1.10 (0.92–1.31) | 3–5 | 425 (38.6) | 1.55 (1.19–2.03)** |
|  |  | ≥4 | 260 (54.5) | 1.46 (1.21–1.77)*** | ≥6 | 376 (62.8) | 2.39 (1.80–3.18)*** |
|  | - AHF cohort: HFpEF | 0–1 | 101 (38.8) | 1.00 (reference) | 0–2 | 41 (22.2) | 1.00 (reference) |
|  |  | 2–3 | 185 (41.8) | 0.83 (0.65–1.06) | 3–5 | 214 (42.8) | 1.85 (1.32–2.59)*** |
|  |  | ≥4 | 93 (43.3) | 0.74 (0.55–0.99)* | ≥6 | 124 (53.2) | 1.88 (1.30–2.72)*** |
| HF hospitalization | HFrEF-ARNI cohort | 0–1 | 94 (14.4) | 1.00 (reference) | 0–2 | 62 (11.7) | 1.00 (reference) |
|  |  | 2–3 | 102 (18.1) | 1.25 (0.94–1.65) | 3–5 | 138 (19.9) | 1.36 (0.99–1.87) |
|  |  | ≥4 | 49 (24.3) | 1.45 (1.02–2.06)* | ≥6 | 45 (23.0) | 1.55 (1.01–2.36)* |
|  | AHF cohort: overall | 0–1 | 152 (17.8) | 1.00 (reference) | 0–2 | 80 (14.5) | 1.00 (reference) |
|  |  | 2–3 | 321 (22.3) | 1.20 (0.98–1.46) | 3–5 | 354 (22.1) | 1.55 (1.21–1.99)*** |
|  |  | ≥4 | 135 (19.5) | 1.11 (0.88–1.41) | ≥6 | 174 (20.9) | 1.76 (1.32–2.36)*** |
|  | - AHF cohort: HFrEF | 0–1 | 104 (17.6) | 1.00 (reference) | 0–2 | 48 (13.1) | 1.00 (reference) |
|  |  | 2–3 | 223 (22.3) | 1.24 (0.98–1.57) | 3–5 | 251 (22.8) | 1.69 (1.23–2.33)** |
|  |  | ≥4 | 95 (19.9) | 1.21 (0.91–1.61) | ≥6 | 123 (20.5) | 1.84 (1.27–2.65)** |
|  | - AHF cohort: HFpEF | 0–1 | 48 (18.5) | 1.00 (reference) | 0–2 | 32 (17.3) | 1.00 (reference) |
|  |  | 2–3 | 98 (22.1) | 1.08 (0.76–1.53) | 3–5 | 103 (20.6) | 1.35 (0.90–2.03) |
|  |  | ≥4 | 40 (18.7) | 0.89 (0.58–1.37) | ≥6 | 51 (21.9) | 1.64 (1.02–2.62)* |

Cutoffs were applied identically to both cohorts. Hazard ratios are relative to the lowest category and were adjusted for age, sex, LVGLS, LASr, and natriuretic peptide level (decile). Each outcome analysis included patients with valid follow-up information for the respective outcome. HFrEF was defined as LVEF <50% and HFpEF as LVEF ≥50%. \*P <0.05; \*\*P <0.01; \*\*\*P <0.001.
Abbreviations: AHF; acute heart failure; ARNI, angiotensin-receptor-neprilysin inhibitor; CI, confidence interval; CKM, cardiovascular-kidney-metabolic; HF, heart failure; HFpEF, heart failure with preserved ejection fraction; HFrEF, heart failure with reduced ejection fraction; HR, hazard ratio; LASr, left atrial reservoir strain; LVEF, left ventricular ejection fraction; LVGLS, left ventricular global longitudinal strain.

**Supplemental Table 4.** Association of CKM burden with HF hospitalization accounting for the competing risk of death.

| Cohort | HF hospitalization N (%) | Death before HF hospitalization N (%) | Binary CKM burden score |  | Graded CKM burden score |  |
|---|---|---|---|---|---|---|
|  |  |  | Cause-specific HR | Fine–Gray sHR | Cause-specific HR | Fine–Gray sHR |
| HFrEF-ARNI cohort | 245 (17.3) | 80 (5.6) | 1.11 (1.02–1.21)* | 1.10 (1.00–1.20)* | 1.10 (1.02–1.19)* | 1.09 (1.01–1.18)* |
| AHF cohort: overall | 608 (20.4) | 968 (32.5) | 1.03 (0.97–1.09) | 1.02 (0.97–1.08) | 1.08 (1.03–1.14)** | 1.04 (1.00–1.10) |
| - AHF cohort: HFrEF | 422 (20.4) | 670 (32.4) | 1.06 (0.98–1.13) | 1.04 (0.97–1.11) | 1.08 (1.02–1.15)* | 1.03 (0.98–1.10) |
| - AHF cohort: HFpEF | 186 (20.3) | 298 (32.5) | 0.97 (0.88–1.08) | 1.00 (0.90–1.10) | 1.08 (0.99–1.17) | 1.06 (0.97–1.15) |

HRs and sHRs are presented per 1-point increment in the score and were adjusted for age, sex, LVGLS, LASr, and natriuretic peptide level (decile). Each outcome analysis included patients with valid follow-up information for the respective outcome. Cause-specific HRs were estimated using Cox models censoring at death. Fine–Gray sHRs were estimated with death before HF hospitalization as the competing event, by maximizing the weighted partial likelihood with inverse-probability-of-censoring weights and a robust sandwich variance estimator. Competing deaths were defined as deaths occurring before any HF hospitalization. HFrEF was defined as LVEF <50% and HFpEF as LVEF ≥50%. \*P <0.05; \*\*P <0.01; \*\*\*P <0.001.
Abbreviations: AHF, acute heart failure; ARNI, angiotensin-receptor-neprilysin inhibitor; CKM, cardiovascular-kidney-metabolic; HF, heart failure; HFpEF, heart failure with preserved ejection fraction; HFrEF, heart failure with reduced ejection fraction; HR, hazard ratio; LASr, left atrial reservoir strain; LVEF, left ventricular ejection fraction; LVGLS, left ventricular global longitudinal strain; sHR, subdistribution hazard ratio.

**Supplemental Table 5.** Incremental prognostic value of the binary and graded CKM burden scores for all-cause death and HF hospitalization.

| Outcome | Cohort | N | Events,<br>N (%) | Baseline model<br>χ<sup>2</sup> | Binary score<br>Δχ<sup>2</sup> | Graded score<br>Δχ<sup>2</sup> | C-index<br>binary → graded | ΔC-index (95% CI) |
|---|---|---|---|---|---|---|---|---|
| All-cause death | HFrEF-ARNI cohort | 1,419 | 128 (9.0) | 118.3 | 2.6 | 7.9** | 0.615 → 0.699 | +0.085<br>(+0.043 to +0.124)*** |
|  | AHF cohort: overall | 2,984 | 1244 (41.7) | 593.1 | 5.5* | 62.5*** | 0.546 → 0.626 | +0.080<br>(+0.066 to +0.093)*** |
|  | - AHF cohort: HFrEF | 2,066 | 865 (41.9) | 474.7 | 14.9*** | 55.3*** | 0.572 → 0.643 | +0.072<br>(+0.057 to +0.087)*** |
|  | - AHF cohort: HFpEF | 918 | 379 (41.3) | 138.0 | 3.3 | 10.1** | 0.485 → 0.583 | +0.098<br>(+0.073 to +0.122)*** |
| HF hospitalization | HFrEF-ARNI cohort | 1,418 | 245 (17.3) | 97.3 | 5.3* | 6.3* | 0.571 → 0.606 | +0.035<br>(+0.002 to +0.065)* |
|  | AHF cohort: overall | 2,984 | 608 (20.4) | 90.1 | 1.1 | 10.2** | 0.522 → 0.564 | +0.042<br>(+0.024 to +0.060)*** |
|  | - AHF cohort: HFrEF | 2,067 | 422 (20.4) | 77.5 | 2.3 | 6.6* | 0.537 → 0.571 | +0.034<br>(+0.012 to +0.056)** |
|  | - AHF cohort: HFpEF | 917 | 186 (20.3) | 23.8 | 0.3 | 3.2 | 0.487 → 0.546 | +0.059<br>(+0.024 to +0.094)*** |

The baseline model included age, sex, LVGLS, LASr, and natriuretic peptide level (decile). Δχ<sup>2</sup> is the increase in the global likelihood-ratio χ<sup>2</sup> when each CKM burden score is added to the baseline model, tested with 1 degree of freedom. C-indices were calculated using each CKM burden score alone, without covariate adjustment; ΔC-index represents the graded minus binary score, with 95% CIs derived from paired bootstrap resampling (1,000 replicates, fixed random seed). P values were derived from likelihood-ratio tests for Δχ<sup>2</sup> and normal approximation tests using bootstrap standard errors for ΔC-index. \*P <0.05; \*\*P <0.01; \*\*\*P <0.001
Each outcome analysis included patients with valid follow-up information for the respective outcome. HFrEF was defined as LVEF <50% and HFpEF as LVEF ≥50%.
Abbreviations: AHF, acute heart failure; ARNI, angiotensin-receptor-neprilysin inhibitor; CI, confidence interval; CKM, cardiovascular-kidney-metabolic; HF, heart failure; HFpEF, heart failure with preserved ejection fraction; HFrEF, heart failure with reduced ejection fraction; LASr, left atrial reservoir strain; LVEF, left ventricular ejection fraction; LVGLS, left ventricular global longitudinal strain.

## Supplemental Figures

**Supplemental Figure 1.**
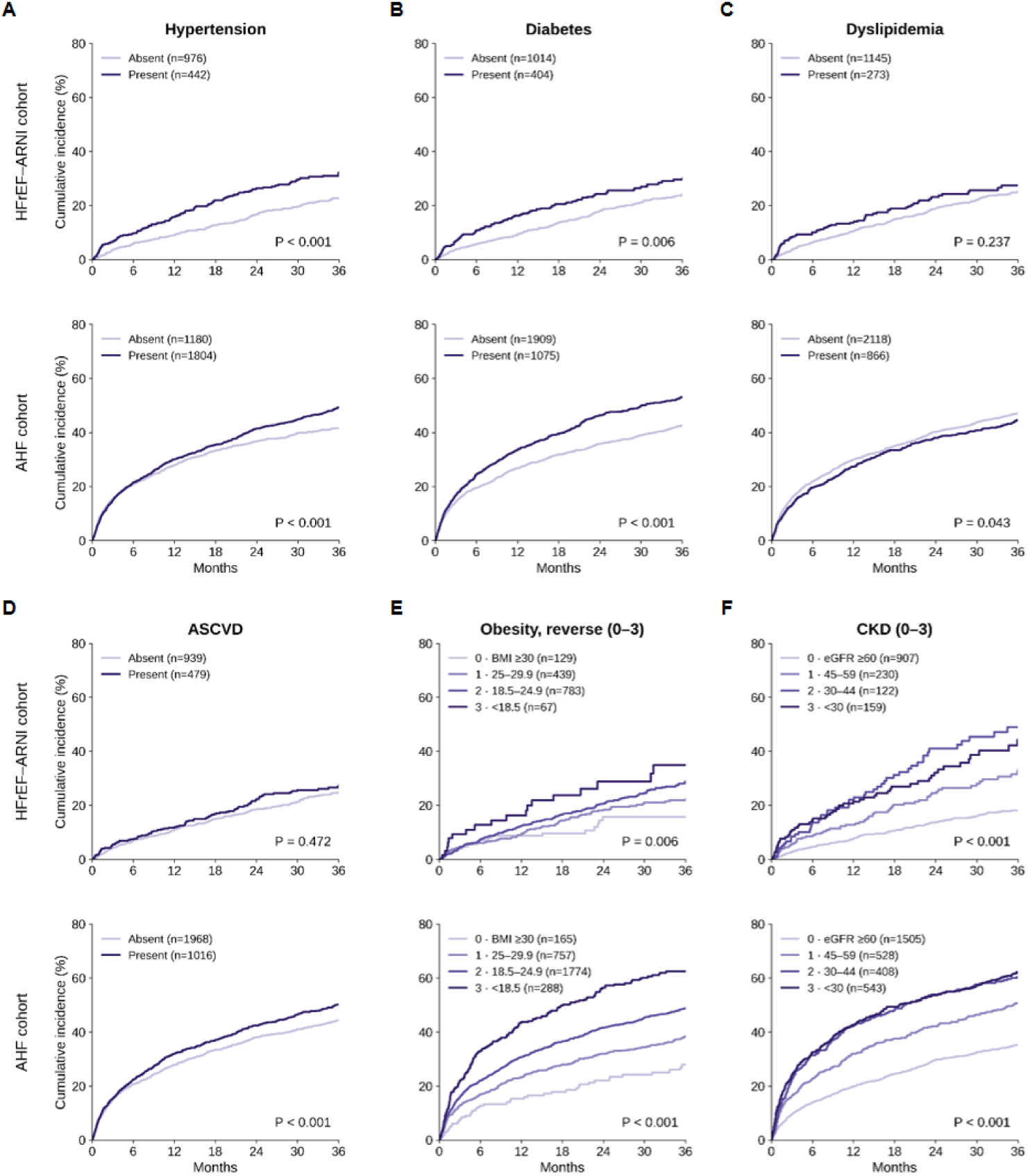
Kaplan–Meier Estimates of the Composite Outcome by Individual CKM Component. (A) Hypertension; (B) diabetes mellitus; (C) dyslipidemia; (D) ASCVD; (E) obesity, reverse; and (F) CKD. Within each panel, the upper plot shows the HFrEF-ARNI cohort (n = 1,418) and the lower plot shows the AHF cohort (n = 2,984). Unadjusted Kaplan–Meier estimates of the composite outcome are shown according to each CKM component. P values are from log-rank tests, and the y-axis scale is common to all panels. Obesity and CKD are shown in the four levels used in the graded score, with obesity reverse-scored so that lower body mass receives a higher score. Abbreviations: AHF, acute heart failure; ARNI, angiotensin-receptor-neprilysin inhibitor; ASCVD, atherosclerotic cardiovascular disease; CKD, chronic kidney disease; CKM, cardiovascular-kidney-metabolic; HFrEF, heart failure with reduced ejection fraction.

**Supplemental Figure 2.**
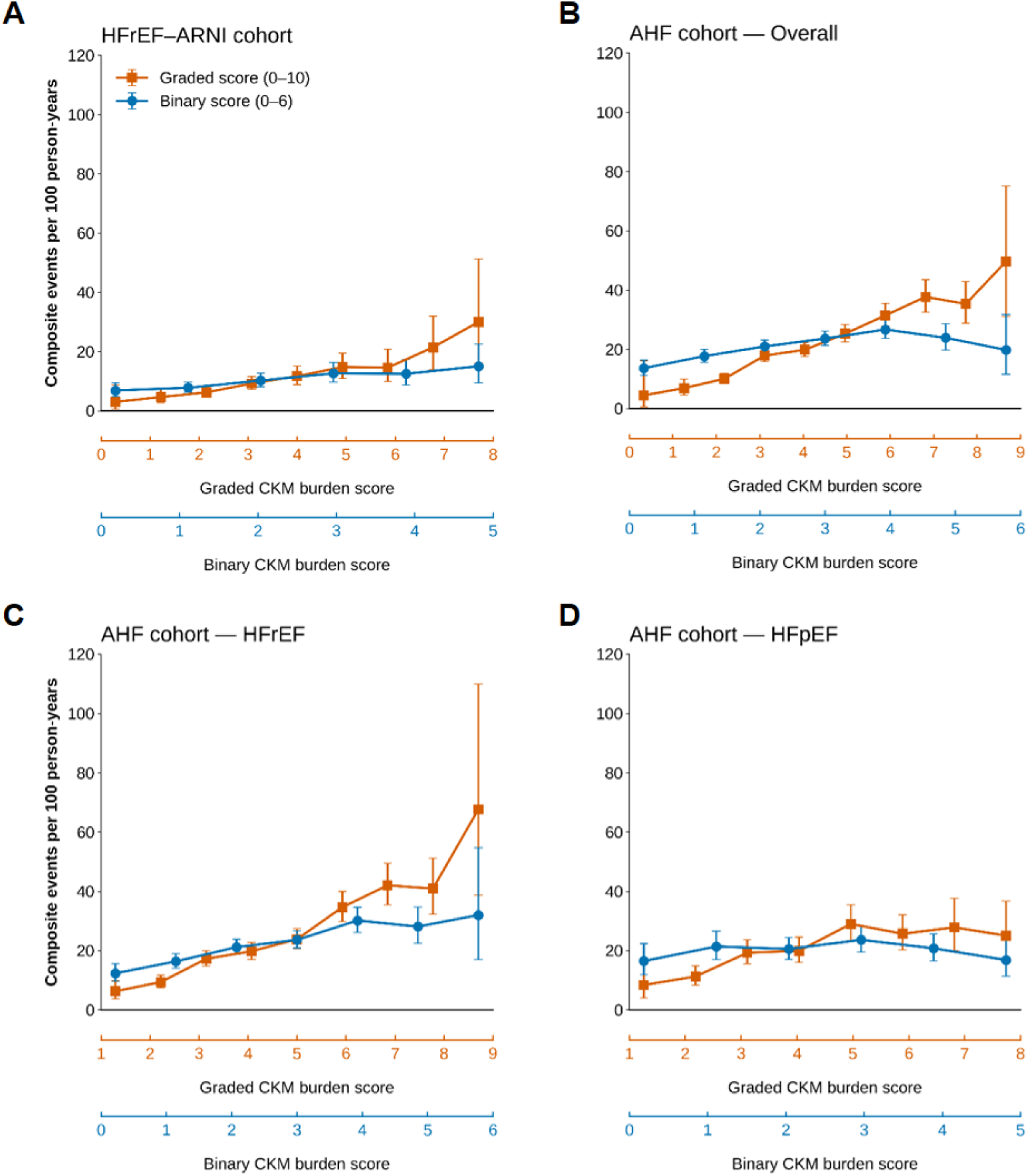
Incidence Rate of the Composite Outcome across CKM Burden Score Levels. (A) HFrEF-ARNI cohort (n = 1,418); (B) AHF cohort **–** Overall (n = 2,984); (C) AHF cohort **–**HFrEF (n = 2,067); and (D) AHF cohort **–** HFpEF (n = 917). Unadjusted incidence rates are shown as events per 100 person-years over the full available follow-up, with exact Poisson 95% confidence intervals. The graded (orange) and binary (blue) scores are shown on separate x-axes spanning their respective observed ranges. Score levels with fewer than 15 patients are not shown. HFrEF and HFpEF were defined as LVEF <50% and ≥50%, respectively. Abbreviations: AHF, acute heart failure; ARNI, angiotensin-receptor-neprilysin inhibitor; CKM, cardiovascular-kidney-metabolic; HFpEF, heart failure with preserved ejection fraction; HFrEF, heart failure with reduced ejection fraction; LVEF, left ventricular ejection fraction.

**Supplemental Figure 3.**
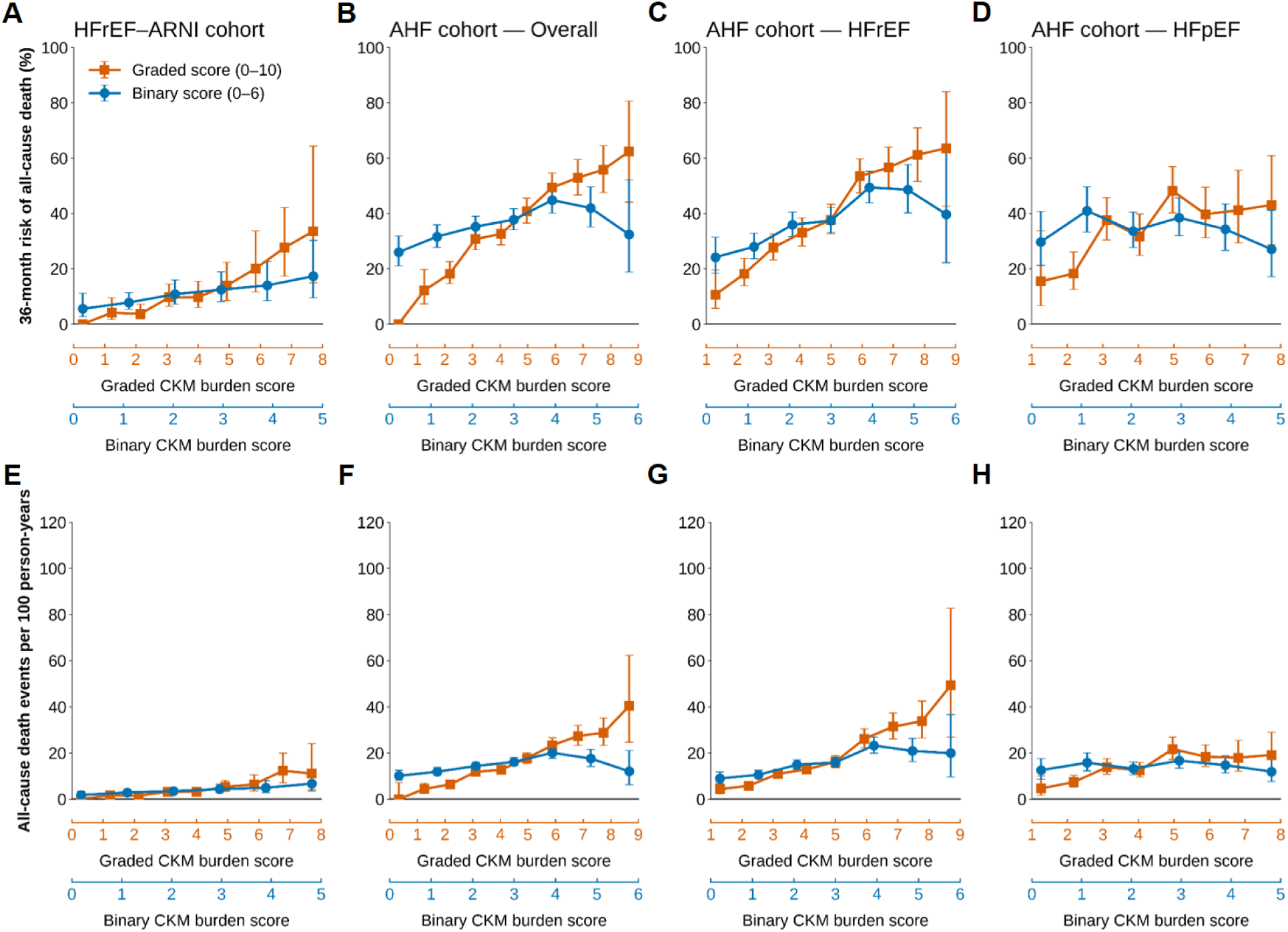
Three-Year Risk and Incidence Rates of All-Cause Death across CKM Burden Score Levels. (A, E) HFrEF-ARNI cohort (n = 1,419); (B, F) AHF cohort **–** Overall (n = 2,984); (C, G) AHF cohort **–** HFrEF (n = 2,066); and (D, H) AHF cohort **–** HFpEF (n = 918). The upper row shows unadjusted Kaplan–Meier estimates of 36-month risk with 95% confidence intervals; the lower row shows incidence rates per 100 person-years over the full available follow-up, with exact Poisson 95% confidence intervals. The y-axis scales differ between rows. The graded (orange) and binary (blue) scores are shown on separate x-axes spanning their respective observed ranges. Score levels with fewer than 15 patients are not shown. HFrEF and HFpEF were defined as LVEF <50% and ≥50%, respectively. Abbreviations: AHF, acute heart failure; ARNI, angiotensin-receptor-neprilysin inhibitor; CKM, cardiovascular-kidney-metabolic; HFpEF, heart failure with preserved ejection fraction; HFrEF, heart failure with reduced ejection fraction; LVEF, left ventricular ejection fraction.

**Supplemental Figure 4.**
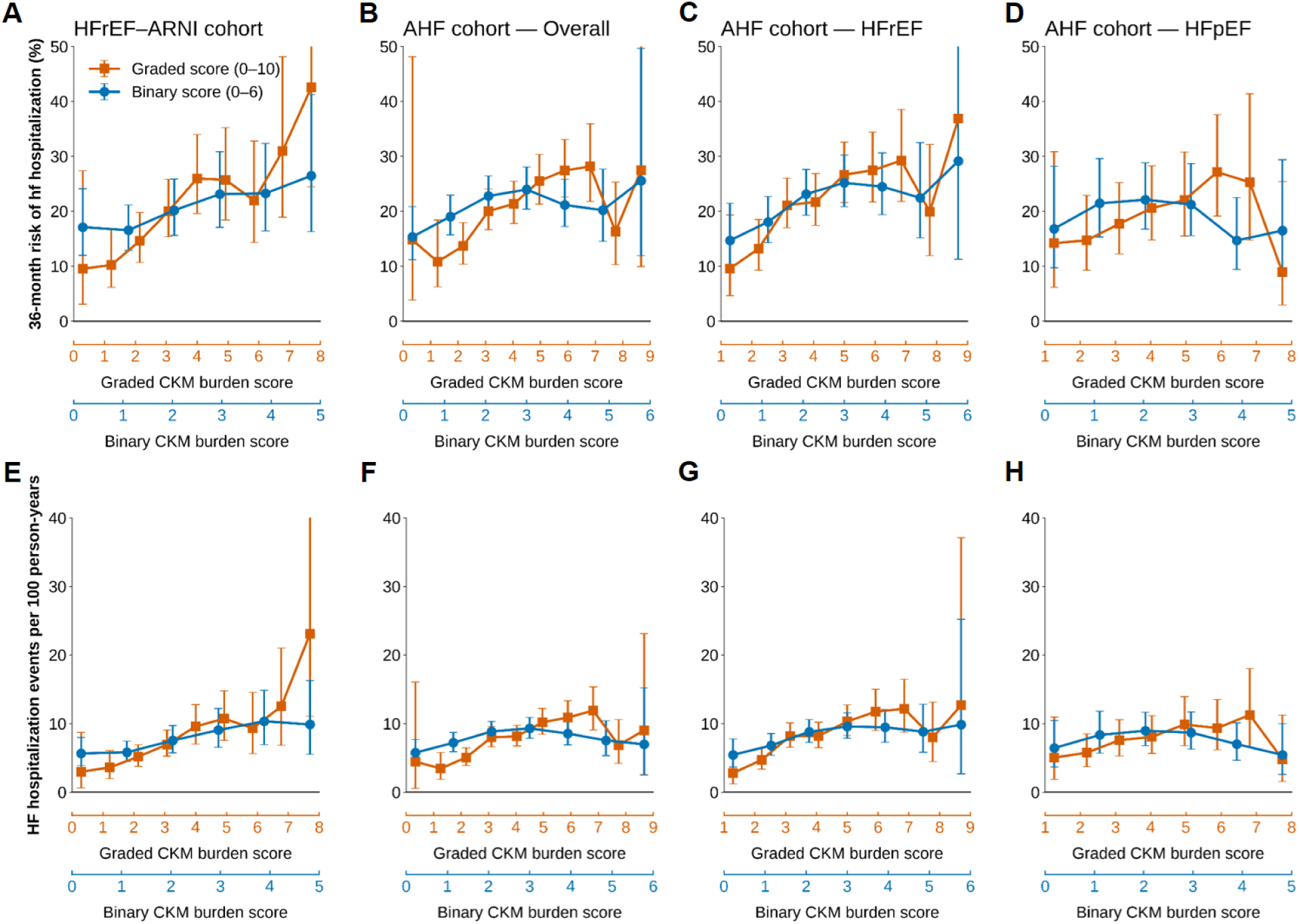
Three-Year Risk and Incidence Rates of HF Hospitalization across CKM Burden Score Levels. (A, E) HFrEF-ARNI cohort (n = 1,418); (B, F) AHF cohort **–** Overall (n = 2,984); (C, G) AHF cohort **–** HFrEF (n = 2,067); and (D, H) AHF cohort **–** HFpEF (n = 917). The upper row shows unadjusted Kaplan–Meier estimates of 36-month risk with 95% confidence intervals; the lower row shows incidence rates per 100 person-years over the full available follow-up, with exact Poisson 95% confidence intervals. The y-axis scales differ between rows. The graded (orange) and binary (blue) scores are shown on separate x-axes spanning their respective observed ranges. Score levels with fewer than 15 patients are not shown. HFrEF and HFpEF were defined as LVEF <50% and ≥50%, respectively. Abbreviations: AHF, acute heart failure; ARNI, angiotensin-receptor-neprilysin inhibitor; CKM, cardiovascular-kidney-metabolic; HF, heart failure; HFpEF, heart failure with preserved ejection fraction; HFrEF, heart failure with reduced ejection fraction; LVEF, left ventricular ejection fraction.

**Supplemental Figure 5.**
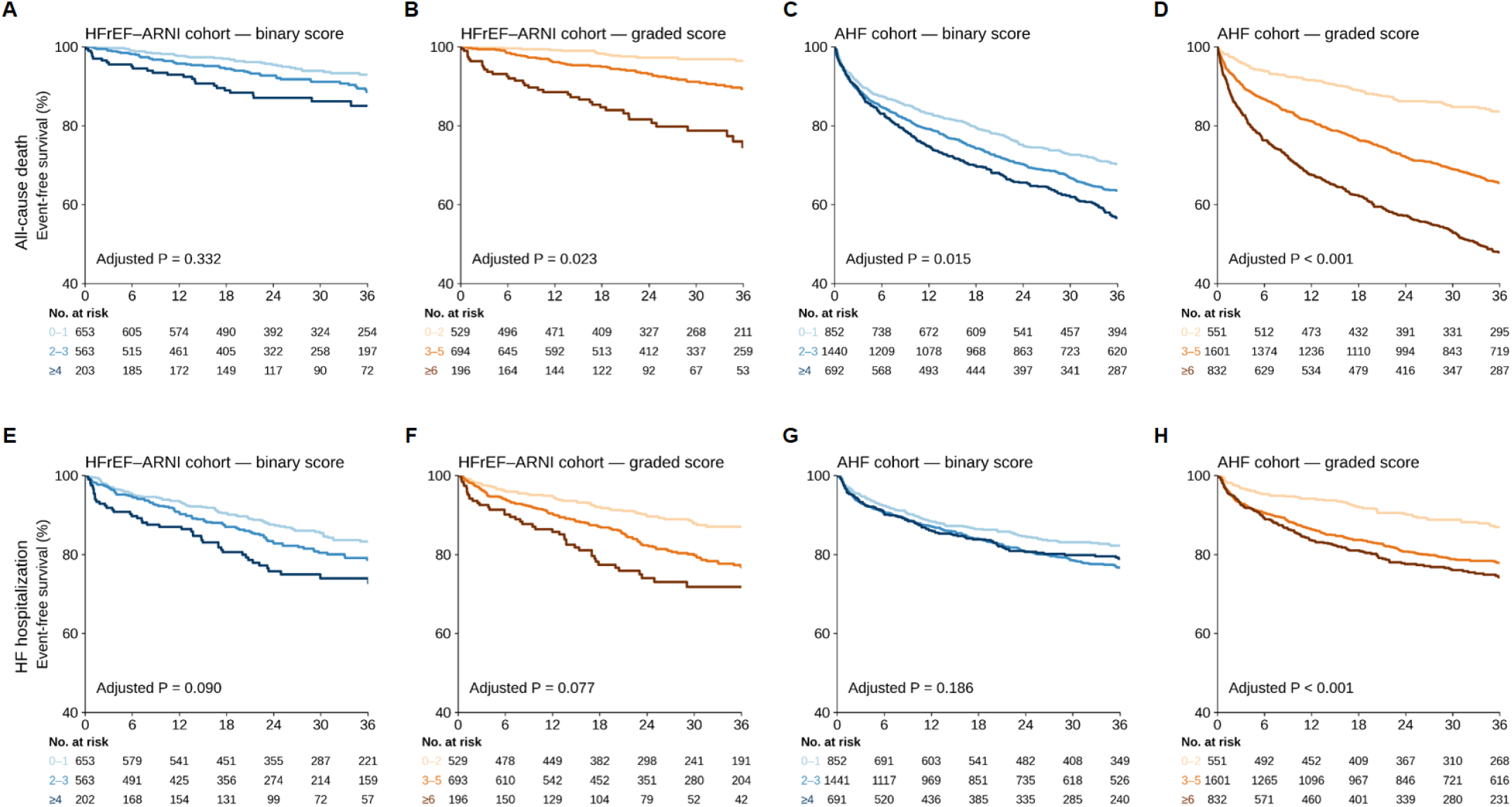
Kaplan–Meier Estimates of All-Cause Death and HF Hospitalization by CKM Burden Score Category. (A, B) All-cause death in the HFrEF-ARNI cohort (n = 1,419); (C, D) all-cause death in the AHF cohort (n = 2,984); (E, F) HF hospitalization in the HFrEF-ARNI cohort (n = 1,418); and (G, H) HF hospitalization in the AHF cohort (n = 2,984). Panels A, C, E, and G show the binary score, and panels B, D, F, and H show the graded score. Unadjusted Kaplan–Meier estimates are shown by CKM burden score category, with numbers at risk below each panel. P values are from Cox models adjusted for age, sex, LVGLS, LASr, and natriuretic peptide level (decile). Binary score categories were 0–1, 2–3, and ≥4; graded score categories were 0–2, 3–5, and ≥6. The y-axis is restricted to 40%–100% to display the curves. Abbreviations: AHF, acute heart failure; ARNI, angiotensin-receptor-neprilysin inhibitor; CKM, cardiovascular-kidney-metabolic; HF, heart failure; HFrEF, heart failure with reduced ejection fraction; LASr, left atrial reservoir strain; LVGLS, left ventricular global longitudinal strain.

**Supplemental Figure 6.**
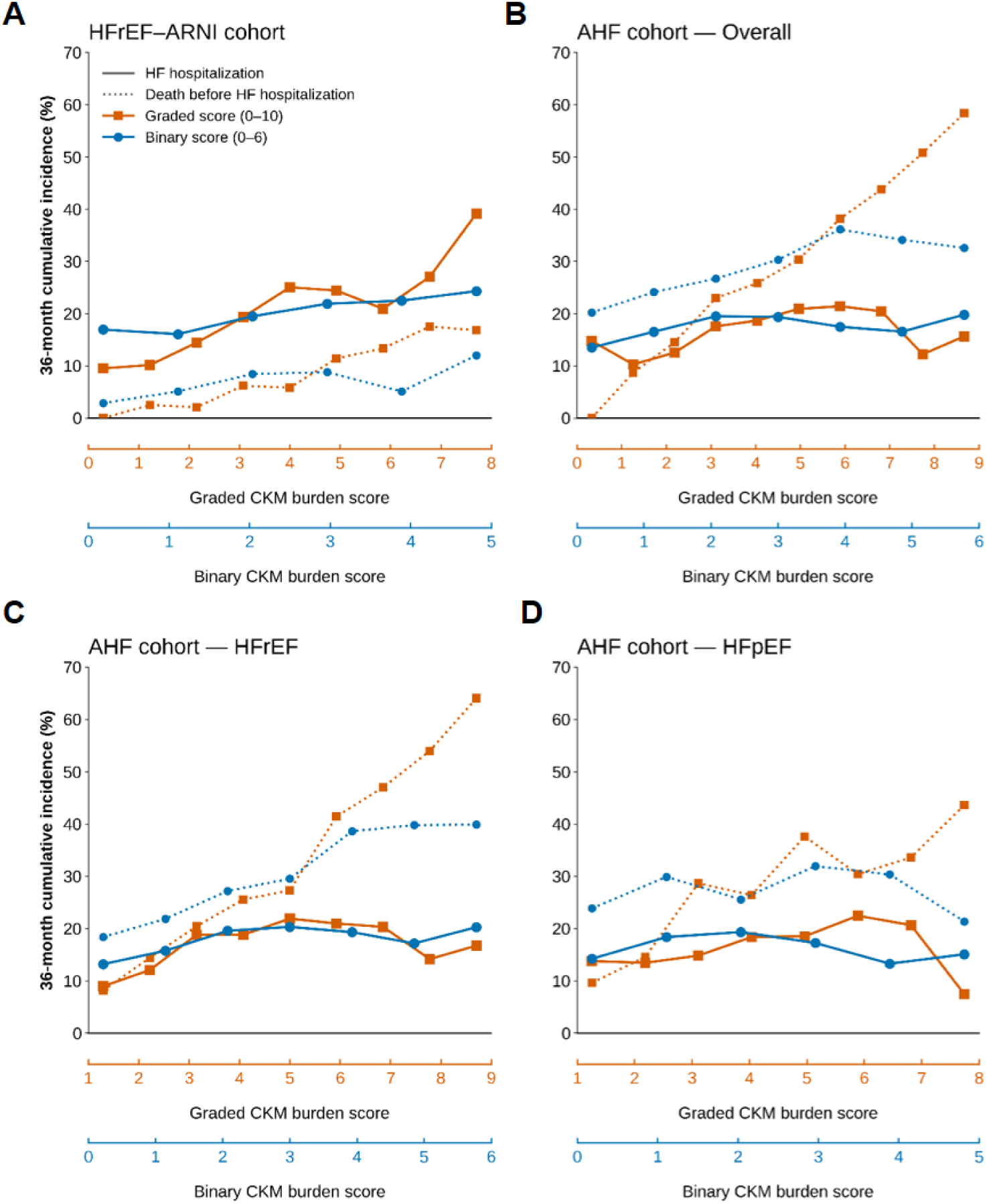
Cumulative Incidence of HF Hospitalization and Death Before HF Hospitalization across CKM Burden Score Levels. (A) HFrEF-ARNI cohort (n = 1,418); (B) AHF cohort **–** Overall (n = 2,983); (C) AHF cohort **–** HFrEF (n = 2,066); and (D) AHF cohort **–** HFpEF (n = 917). Solid and dotted lines show the unadjusted 36-month Aalen–Johansen cumulative incidence of HF hospitalization and death before HF hospitalization, respectively. The graded (orange) and binary (blue) scores are shown on separate x-axes spanning their respective observed ranges. Score levels with fewer than 15 patients are not shown. HFrEF and HFpEF were defined as LVEF <50% and ≥50%, respectively. Abbreviations: AHF, acute heart failure; ARNI, angiotensin-receptor-neprilysin inhibitor; CKM, cardiovascular-kidney-metabolic; HF, heart failure; HFpEF, heart failure with preserved ejection fraction; HFrEF, heart failure with reduced ejection fraction; LVEF, left ventricular ejection fraction.

